# Applying Machine Learning on UK Biobank biomarker data empowers case-control discovery yield

**DOI:** 10.1101/2023.10.10.23296832

**Authors:** Manik Garg, Marcin Karpinski, Dorota Matelska, Lawrence Middleton, Jonathan Mitchell, Amanda O’Neill, Quanli Wang, Andrew Harper, Ryan S. Dhindsa, Slavé Petrovski, Dimitrios Vitsios

**Affiliations:** Centre for Genomics Research, Discovery Sciences, BioPharmaceuticals R&D, AstraZeneca, Cambridge, UK; Centre for Genomics Research, Discovery Sciences, BioPharmaceuticals R&D, AstraZeneca, Waltham, MA, USA; Department of Medicine, University of Melbourne, Austin Health, Melbourne, Victoria, Australia

**Author notes:** These authors contributed equally (Manik Garg, Marcin Karpinski). To whom correspondence should be addressed. &.

## Abstract

Missing or inaccurate diagnoses in biobank datasets can reduce the power of human genetic association studies. We present a machine-learning framework (MILTON) that utilizes the wealth of phenotypic information available in a biobank dataset to identify undiagnosed individuals within the cohort who have biomarker profiles similar to those of positively diagnosed cases. We applied MILTON to perform an augmented phenome-wide association study (PheWAS) based on 405,703 whole exome sequencing samples from UK Biobank, resulting in improved signals for known (p<1×10^−8^) gene-disease relationships alongside 206 novel gene-disease relationships that only achieved genome-wide significance upon using MILTON. To further validate these putatively novel discoveries, we adopt two orthogonal machine learning methods that prioritise gene-disease relationships using comprehensive publicly available datasets alongside a biological insights knowledge graph. For additional clinical translation utility, MILTON outputs a disease-specific biomarker set per disease as well as comorbidity clusters across ICD10 disease codes based on shared biomarker profiles of positively labelled cases. All the extracted associations and biomarker importance results for the 3,308 studied binary traits will be made available via an interactive web-portal.

## Introduction

Understanding the role of genetic variation in human disease is increasingly recognized as a key component in successful drug development. Historically, most association studies used phenotype-specific cohorts, in which cases and controls were recruited to a study in a disease-specific manner. More recently, large-scale biobanks with linked genetic and phenotypic data have emerged as a vital source for studying a wide range of diseases. Rather than focusing on one specific disease, these resources enable “phenome-wide association studies” (PheWAS)^1^, in which genetic variation is systematically tested for association with all available phenotype data. One limitation, however, is that many of the phenotype definitions in biobank-level analyses rely on billing codes and self-reporting, introducing potential misclassification of participants, missing data, and variability in defining case-control cohrots^1^. Identifying individuals who may, in fact, be cases who are either not coded properly in the resource or have not yet been clinically diagnosed (i.e., “cryptic cases”) remains an open challenge.

The UK Biobank (UKB; https://www.ukbiobank.ac.uk) is one of the largest biobank cohorts, including health record and genetic sequencing data from half a million individuals aged between 40 and 69 at recruitment. There is a rich catalogue of phenotype data for each participant in addition to continuously updated health record data, biometric measurements, lifestyle indicators, blood and urine biomarkers, and imaging. We and others have performed PheWAS on UKB data, which have revealed a variety of new genetic associations and candidate therapeutic targets^2–4^. However, missing data are also common among the UK Biobank phenotypes, especially for individuals who have only been seen in outpatient settings. Augmented phenotyping would enable better case-control cohort definitions and increase statistical power for gene discovery within existing sample sizes.

There have been efforts to manually identify cryptic cases within the UKB for isolated diseases based on imaging and biomarker data. While these clinical risk algorithms have been successful for several phenotypes ^5–9^, it is less feasible or scalable to manually define similar algorithms across the entire phenome. Other approaches use both biomarkers and single nucleotide polymoprhisms (SNPs) to predict disease risk^10^. However, using genetic data to augment case cohorts introduces the risk of circularity and bias for downstream genetic association tests on those cohorts.

Here, we introduce a systematic method for defining augmented case definitions using commonly measured clinical biomarkers and other quantitative traits to maximize the utility of case-control cohort-level resources. Our machine-learning approach, MILTON (MachIne Learning with phenoType associatONs), uses 67 UK Biobank quantitative measurements, including sex and age as covariates, to impute missing or undiagnosed phenotypes and in doing so constructing augmented case-control cohort definitions to complement the baseline definitions. Using these predictions, we repeated a phenome-wide association study on 3,308 phenotypes in 405,703 UK Biobank exomes, identifying several putative novel gene-phenotype associations that did not achieve significance in the baseline PheWAS from the same test cohort.

## Results

### Overview

Clinical biomarkers play a key role in diagnosing many common chronic diseases, as they provide measurable indications of the presence or severity of a condition. In the setting of phenome-wide association studies, they also provide an opportunity to identify cryptic or misclassified cases. Here, we introduce a machine learning method, MILTON, that uses quantitative biomarkers as features to predict augmented case status for 3,308 disease phenotypes (**Fig. 1**; see Methods). While the MILTON framework could be applied to any biobank-level dataset that includes biomarker data, we apply it here to the UK Biobank (UKB), a collection of 405,703 high quality, predominantly unrelated, and of European ancestry exome sequencing samples with linked health record data.

**Figure 1.**
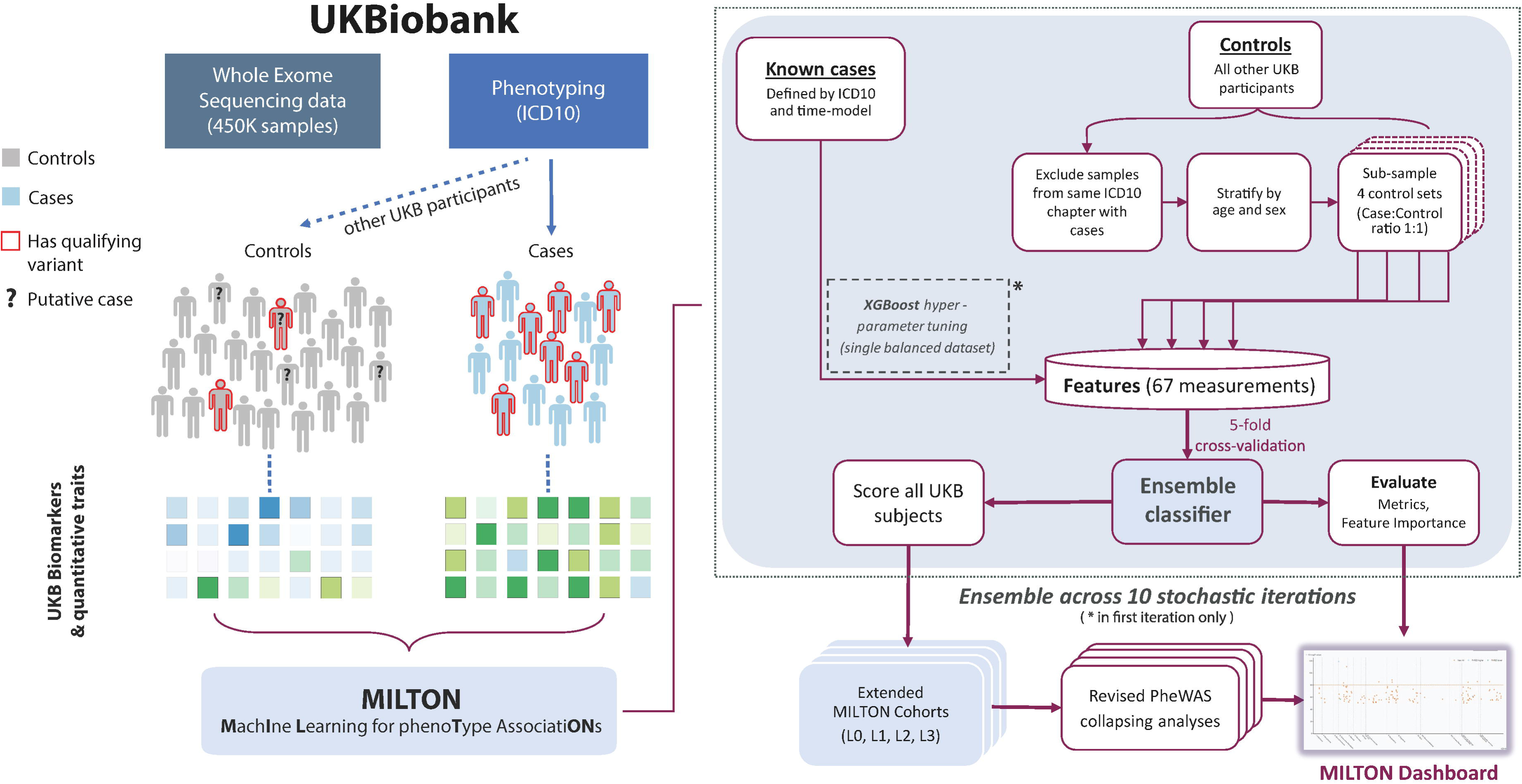
MILTON flowchart. Individuals diagnosed with certain ICD10 in UK Biobank are herein referred to as “cases” and all remaining individuals as “controls” for that ICD10. Both cases and controls can have qualifying variants (QVs), such as protein truncating variants, in a given gene. The objective of rare-variant collapsing analysis is to identify QVs in which gene are enriched in either cases or controls. Some controls may not yet be diagnosed with a given ICD10 or are incorrectly classified. MILTON aims to identify these individuals by checking if they share similar biomarker profile to known cases (represented by the shades of green). The predicted cases are eventually merged with the known cases cohort along with a revised control set to perform an updated PheWAS collapsing analysis.

In MILTON, we first select cases and controls for each ICD10 code^11^ (Data-Coding 19), and then extract biomarker values for each participant to infer a disease-specific signature. Next, we use an extreme gradient boosting (XGBoost^12^) model on a single balanced case-control set to find the optimal hyperparameters for a given ICD10 code. We train an ensemble model with 5-fold cross-validation employing up to four balanced case-control sets (bound defined by the initial number of cases and total UKB cohort size) and repeat for 10 stochastic iterations to ensure the entire control set is fed to the model. We then apply the final model to the entire UKB cohort to predict the probability of each UKB participant being a case for that given disease. We then assign individuals as “cases” based on four different probability thresholds (named L0-L3; see Methods). The L0 class includes the strictest cut-off whereas L3 uses the most lenient, including the largest number of predicted cases. Finally, we repeat rare-variant collapsing analysis^2^ on each of the augmented cohorts and compare the results against the baseline cohorts used to train the models, as well as against the original PheWAS results that relied exclusively on positive labelled cases^2^.

### Defining optimal time-lag between sample collection and diagnosis date

In the UKB, samples for biomarker measurement may have been collected as much as 15 years before or 50 years after the corresponding individual was diagnosed with a disease (Figure 2A). To determine the effect of this time-lag on predictive performance, we trained MILTON models on cases selected according to three different time-models: prognostic, diagnostic, and time-agnostic (Figure 2A) and five different time-lags (see Methods). The prognostic model uses all individuals who received diagnosis up to a cut-off number of years (10 years) after biomarker sample collection, the diagnostic model uses all individuals who received diagnosis up to 10 years before biomarker sample collection, and the time-agnostic model uses all diagnosed individuals for model development (see Methods). The 10 years’ time lag was selected as optimal after a sensitivity analysis on the effect of sample collection and diagnosis time-lag across 400 randomly selected ICD10 codes (see Methods, Supplementary Fig. 1).

**Figure 2.**
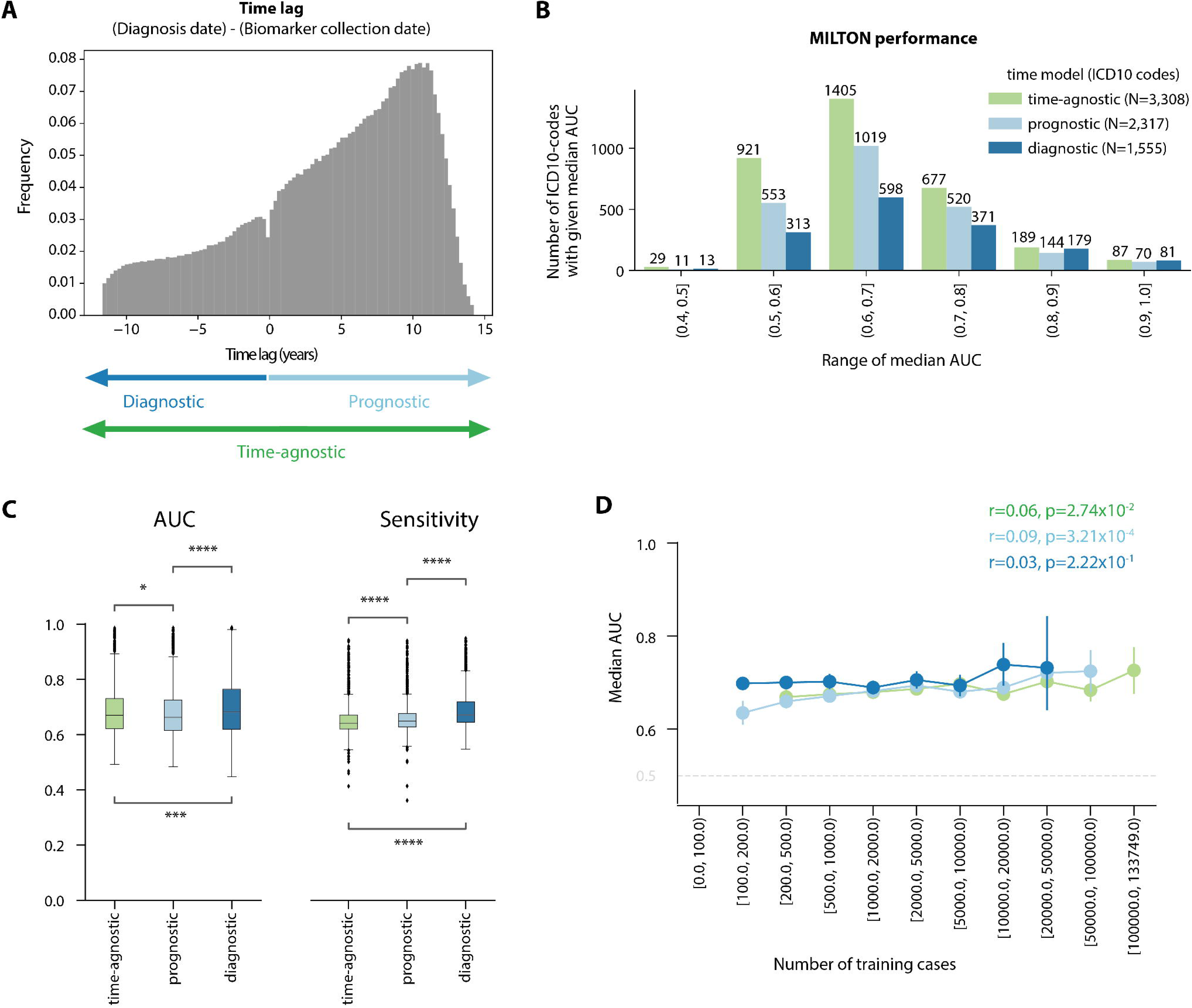
**A** Schematic showing how different time-models are defined and the frequency of individuals that had biomarker sample collection certain years before or after diagnosis date. Earliest diagnosis date recorded in UKB fields 41270, 40001, 40002 or 40006 was taken for each individual (see Methods and Materials). The minimum and maximum lag are trimmed at 0.01 and 0.999 quantile, respectively. **B** Bar-plot showing the distribution of ICD10 codes for which MILTON had median AUC in the given range across ten replicates. **C** Boxplots comparing the median AUC and sensitivity performance of MILTON models across ten replicates trained on 1,357 ICD10 codes under different time-models. Mann-Whitney U test, two-sided p-values are shown. *: 1.00e-02 < p ≤ 5.00e-02; **: 1.00e-03 < p ≤ 1.00e-02; ***: 1.00e-04 < p ≤ 1.00e-03; ****: p ≤ 1.00e-04. Each box-plot shows median as centre line, 25^th^ percentile as lower box limit, 75^th^ percentile as upper box-limit, whiskers extend to 25^th^ percentile – 1.5 * interquartile range at the bottom and 75^th^ percentile + 1.5*interquartile range at the top, points denote outliers. **D** Line-plot showing the distribution of median AUC across ten replicates with increasing number of training cases per ICD10-code across different time-models. Error-bar represents 95% confidence interval. Pearson correlation coefficients (r) and p-values (p) for each time-model are indicated.

### MILTON phenome-wide performance

After running MILTON across the phenome (n=13,942 disease phenotypes) we had results for 3,308, 2,317 and 1,555 ICD10 codes based on time-agnostic, prognostic and diagnostic models, respectively, that satisfied our minimum set of robustness criteria (Supplementary Table 1). Time-agnostic and prognostic models yielded more ICD10 codes than diagnostic models since more individuals have received a diagnosis after biomarker (sample) collection in the UKB (Figure 2A). The distribution of the number of ICD10 codes within each AUC bin is shown in Figure 2B. Specifically, MILTON achieved an AUC>0.7 for 953 ICD10 codes, AUC>0.8 for 276 ICD10 codes, and AUC>0.9 for 87 ICD10 codes under the time-agnostic model. Furthermore, MILTON achieved an AUC>0.6 for more than half of all ICD10 codes per ICD10 chapter across 13 out of 18 chapters and all three time-models (Supplementary Fig. 2A). Similar to the trend seen in Supplementary Fig. 1A, diagnostic models had higher performance than prognostic models across 1,357 ICD10 codes that had results for all three time-models (median AUC_diagnostic_ vs AUC_prognostic_: 0.682 vs 0.663; median Sensitivity_diagnostic_ vs Sensitivity_prognostic_: 0.671 vs 0.649; Figure 2C, Supplementary Data 1A). Overall, as the number of cases available for training per ICD10 increased, AUC remained stable (Figure 2D), specificity had a slightly increasing trend (Pearson’s *r* > 0, p<1×10^−13^), while sensitivity had a slightly decreasing trend (Pearson’s *r* < 0, p<1×10^−8^; Supplementary Fig. 2B).

### MILTON outperforms polygenic risk scores in disease signature inference

We compared the performance of MILTON models trained on 67 quantitative traits (see Methods and Materials) with those trained on either 28 disease-specific standard polygenic risk scores (PRS)^13^, the union of 36 disease-specific or trait-specific standard PRS.

For comparison with 28 disease-specific PRS, we mapped disease-terms to corresponding ICD10 codes using string matching followed by manual curation (Supplementary Table 2). We observed that models trained on 67 quantitative traits significantly outperformed those trained on single disease-specific PRS + sex + age as features for 117 out of 145 ICD10 codes for time-agnostic models (median AUC_67 traits_ vs AUC_disease-specific PRS_: 0.72 vs 0.64, Mann-Whitney U two-sided p=2.39×10^−11^; Table 1, Fig. 3A, Supplementary Fig. 3A, Supplementary Data 2A). This was consistent when models were trained on individuals with European ancestry only (Supplementary Fig. 3B). We also observed the same trend for 98 out of 119 ICD10 codes using prognostic models (median AUC_67 traits_ vs AUC_disease-specific PRS_: 0.73 vs 0.64, Mann-Whitney U two-sided p=6.18×10^−12^; Table 1, Supplementary Fig. 3C, D, Supplementary Data 2B). It is of note that PRS scores for breast cancer (C50) and melanoma (C43, D03) performed better than the 67 quantitative traits in both prognostic and time-agnostic models (Supplementary Fig. 3A, D). This may be partly because the biomarkers used in training MILTON carry less predictive value for these solid cancers. In a separate analysis, we trained MILTON models on all 36 disease-specific or trait-specific standard PRS and again observed that models trained on 67 traits significantly outperformed those trained on PRS for 468 randomly selected ICD10 codes (median AUC_67 traits_ vs AUC_all PRS_: 0.68 vs 0.56, Mann-Whitney U two-sided p=1.13×10^−81^; Table 1, Fig. 3B, Supplementary Data 2C).

**Figure 3.**
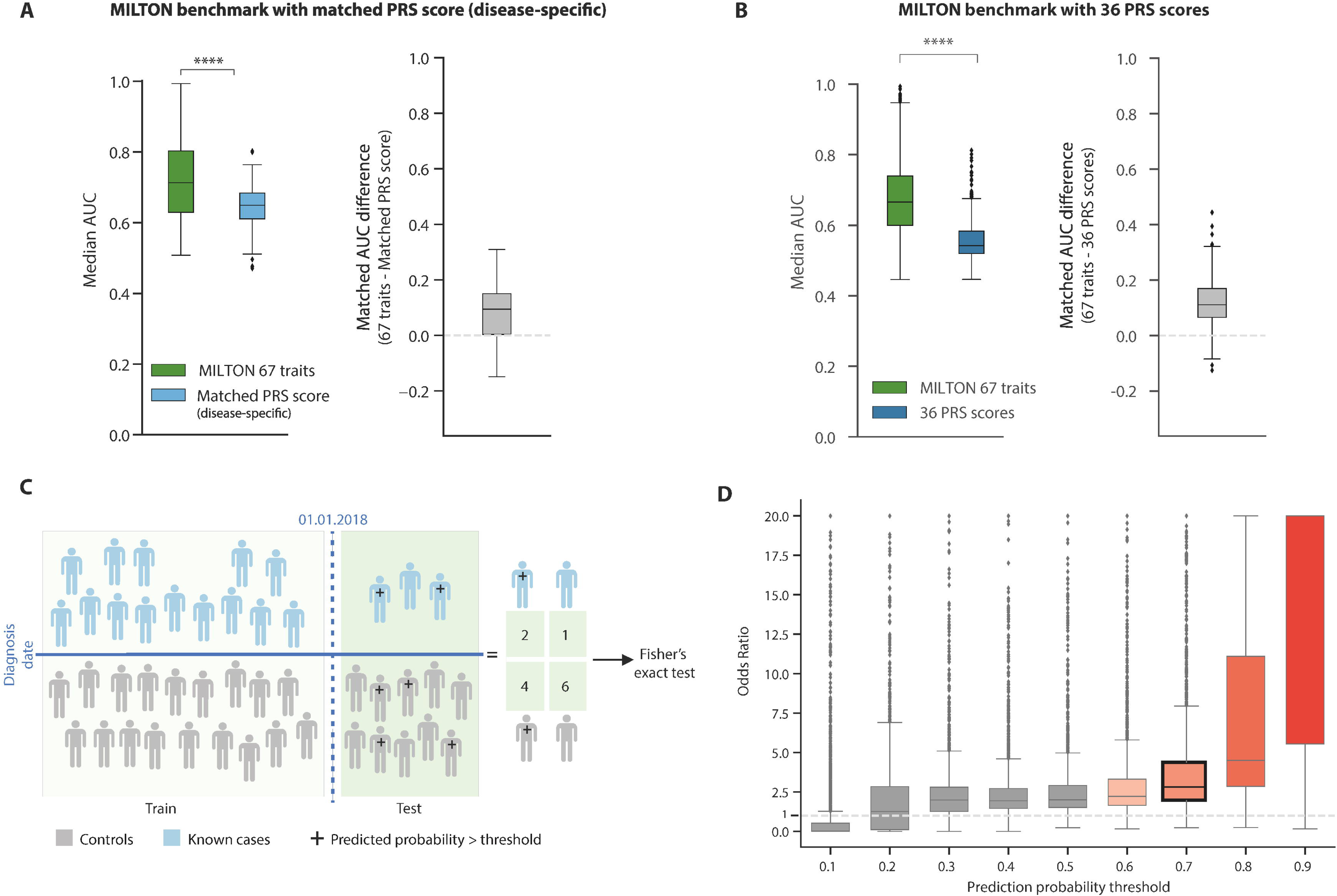
**A** Box plots comparing the performance of MILTON time-agnostic models when trained on 67 traits vs disease-specific PRS scores across 154 ICD10 codes. Right panel: x-axis represents Median AUC_67 traits_ – Median AUC_Matched PRS_ for matched ICD10 codes. **B** Box plots comparing the performance of MILTON time-agnostic models when trained on 67 traits vs all 36 PRS scores across 468 ICD10 codes. **C** Overview of capped analysis. Here, all individuals diagnosed until 01/01/2018 were used during model training and all individuals diagnosed after 01/01/2018 were used as a test set for performance assessment. A 2×2 contingency table was constructed. Top-left: known cases diagnosed after 01/01/2018 and assigned high prediction probability of being a case by MILTON; Top-right: known cases diagnosed after 01/01/2018 and assigned low prediction probability by MILTON; Bottom-left: individuals never diagnosed with given disease and assigned high prediction probability by MILTON; Bottom-right: individuals never diagnosed with given disease and assigned low prediction probability by MILTON. Fisher’s exact test was performed on this 2×2 contingency table to calculate odds ratio and p-value. **D** Distribution of odds ratio obtained from Fisher’s exact test in capped analysis across multiple prediction probability thresholds, indicating MILTON’s power to predict known cases hidden from the training set. In **A, B** and **D**, each box-plot shows median as centre line, 25^th^ percentile as lower box limit, 75^th^ percentile as upper box-limit, whiskers extend to 25^th^ percentile – 1.5 * interquartile range at the bottom and 75^th^ percentile + 1.5*interquartile range at the top, points denote outliers. ****: Mann-Whitney U test, two-sided p-values < 10^−4^; ns: Mann-Whitney U test, two-sided p-values >0.05.

**Table 1.** Summary of MILTON models’ performance when trained on various feature sets and time-models.

| Time-model<br>(abs(time-lag)) | Number<br>of ICD10<br>codes | Features | AUC [0, 1] | Sensitivity | Specificity |
| --- | --- | --- | --- | --- | --- |
| Time-agnostic (no lag) | 145 | Disease-specific PRS + sex + age | $0.64 \pm 0.06$ | $64.85\% \pm 5.06\%$ | $55.50\% \pm 7.80\%$ |
| | | 67 traits | $0.73 \pm 0.12$ | $71.20\% \pm 8.24\%$ | $64.21\% \pm 14.73\%$ |
| Prognostic ($\leq 10$ years) | 119 | Disease-specific PRS + sex + age | $0.64 \pm 0.06$ | $64.10\% \pm 5.98\%$ | $55.34\% \pm 7.37\%$ |
| | | 67 traits | $0.73 \pm 0.12$ | $70.64\% \pm 7.97\%$ | $64.24\% \pm 14.47\%$ |
| Time-agnostic (no lag) | 468 | All 36 PRS + sex + age | $0.56 \pm 0.06$ | $65.23\% \pm 3.21\%$ | $43.04\% \pm 8.04\%$ |
| | | 67 traits | $0.68 \pm 0.10$ | $67.11\% \pm 6.23\%$ | $59.17\% \pm 12.55\%$ |

### MILTON successfully predicts individuals who are subsequently diagnosed with a given ICD10

In order to assess the effectiveness of MILTON in predicting genuine cases, we sought to determine whether individuals assigned a high case probability (0.7 ≤ P_case_ ≤ 1) by MILTON were eventually diagnosed with those ICD10 codes in subsequent phenotype refreshes. To investigate this, we trained MILTON models solely on cases diagnosed prior to January 1, 2018. Subsequently, we analysed the predicted probability scores for cases diagnosed after this date (capped analysis, Figure 3C).

We focused on ICD10 codes with a minimum of 30 individuals diagnosed after January 1, 2018. Among these 2,413 ICD10 codes that met the specified criteria (Supplementary Table 1, Supplementary Data 1B), 1,951 codes (81%) were significantly enriched in participants who had P_case_ >=0.7 and were subsequently diagnosed after January 1, 2018. This observation was supported by an odds ratio greater than 1 (Fisher’s exact test p-value < 0.05) which persisted across prediction probability thresholds ≥ 0.3 (Figure 3D, Supplementary Table 3).

These results further validate MILTONs ability to predict emerging cases from a pool of current controls and, thus, emphasize its value in augmenting existing positive case labels across the biobank cohort.

### MILTON identifies most predictive features per ICD10 and groups related diseases

We observed that MILTON assigned high feature importance scores (FIS) to at least one of the listed biomarkers for the corresponding disease chapter^14^ (Figure 4A, Supplementary Data 3A). For example, glycated haemoglobin (HbA1c) and glucose ranked as the top two features for insulin-dependent diabetes mellitus (E10: AUC_all time-models_ = 0.96 ± 0.02; Figure 4B), among other diseases. Cystatin C, microalbumin in urine and creatinine ranked within top five in acute and chronic renal failure (N17: AUC_all time-models_ = 0.81 ± 0.07; N18: AUC_all time-models_ = 0.89 ± 0.07; Figure 4B). Apart from these disease-biomarker associations^14^, we observed that sex ranked second for predicting chronic renal failure (ICD10: N18)^15–18^ and had greater than average FIS for this phenotype compared to other phenotypes (Supplementary Fig. 4B). This indicates that the MILTON models learn how to distinguish between male- and female-specific cut-offs across the different biomarkers, as in certain diseases reference ranges for some biomarkers may be sex-specific. Overall, though, sex was among the least contributing features across all phenotypes MILTON was run for (Supplementary Fig. 12). As a positive validation, we also confirmed that MILTON ranks as top predictive features, biomarkers known to be associated with certain diseases^19–22^ (Figure 4A-B and Supplementary Fig. 4A).

**Figure 4.**
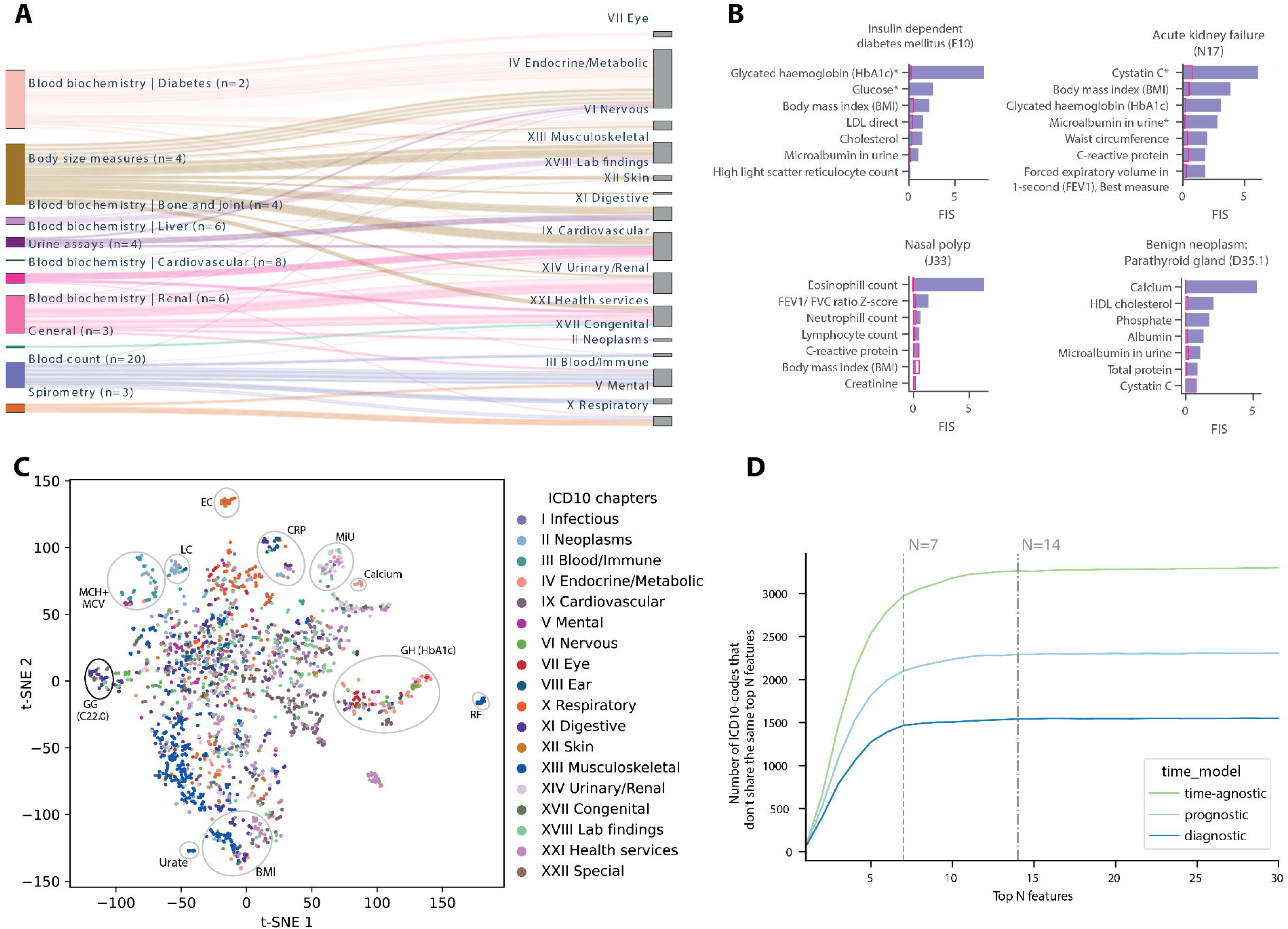
Overview of most important biomarker features learnt by MILTON per ICD10-code. **A** Sankey plot showing mapping from feature categories (left) to ICD10 chapters (right) where each line corresponds to a single feature (with FIS>0.1) to single ICD10-code mapping. Here, n=number of features in each category and ‘|’ denotes the disease area mapped to the given set of biomarkers^14^. Please note that all four biomarkers derived from urine assays were mapped to ‘Renal’ disease area. **B** Bar-plots (in purple) showing top seven features by feature importance scores for four different ICD10 codes (indicated on the top of each bar-plot) under time-agnostic model. Bar-plots in pink show the median FIS of each feature across all ICD10 codes for time-agnostic model. *: represents biomarkers mapped to the given therapy area by clinical experts^14^. E10: Insulin-dependent diabetes mellitus; N17: Acute renal failure; N18: Chronic kidney disease; D35.1: Benign neoplasm of other and unspecified endocrine glands (Parathyroid gland). **C** t-SNE plot where each point corresponds to an ICD10-code, colored by chapter. Cluster highlighted in black oval contains C22.0 ICD10-code. RF=rheumatoid factor, EC=eosinophil count, LC=lymphocyte count, MCV=mean corpuscular volume, MCH=mean corpuscular haemoglobin, GG=gamma glutamyltransferase, CRP=C-reactive protein, MIU=microalbumin in urine, GH = glycated haemoglobin. **D** Line plot showing the number of ICD10 codes that do not share the top N features as a function of N, indicating a quasi-unique biomarker signature per disease, comprised of N>7 features.

We found that ICD10 codes sharing similar biomarker profiles grouped together in a 2-dimensional t-SNE plot (Figure 4C). We zoomed into one cluster containing C22.0 liver cell carcinoma (Figure 4C) to check if individuals diagnosed with this ICD10 code are also diagnosed with other ICD10 codes in the same cluster (see Methods and Materials; Supplementary Data 3B). We observed that 21 out of 35 codes in this cluster were already significantly enriched (p<5×10^−51^) as comorbidities in patients diagnosed with C22.0 (Supplementary Fig. 4C, Supplementary Data 3C).

We also observed that between 7 and 14 features may be sufficient to distinguish one ICD10-code from another, thereby, potentially offering a unique multi-biomarker signature per ICD10-code (Figure 4D). We will be making these top predictive features per disease available in our public portal.

### Disease PheWAS on MILTON augmented case definitions reveal putative novel gene-disease signals

We report MILTON augmented cohorts (L0-L3, Supplementary Fig. 5A, see Methods and Materials) for 2,358 ICD10 codes with AUC>0.6. Here, all MILTON augmented cohorts contain all known cases along with an increasing number of MILTON-predicted cases as we go from L0 (conservative) to L3 (liberal) predictions (see Methods and Materials). Rare-variant collapsing analysis on these augmented cohorts resulted in 2,677 significant gene-ICD10 associations (p_MILTON_<1×10^−8^) between 1,205 ICD10 codes and 149 genes, 99.78% of which are derived by non-synonymous qualifying variant (QVs)^2^ models (Supplementary Data 4). To minimise redundancies, we mapped all associations with sub codes such as N18.0, N18.1 to their 3-character code (N18 in this case). This resulted in 1,323 significant gene-ICD10 associations (p_MILTON_<1×10^−8^) between 454 3-character ICD10 codes, 149 genes with non-synonymous QVs.

To provide a reference dataset, we performed binary PheWAS analysis on the baseline cohort for each ICD10-code and recovered 168 out of 214 gene-ICD10 associations from baseline analyses in the augmented cohorts with same direction of effect (p_MILTON,_ p_Baseline_ <1×10^−8^). We labelled them as ‘known binary’ (Table 2; Supplementary Fig. 5B). Gene-disease associations from ICD10-codes where MILTON had no contribution to case cohort extension are excluded from the reported baseline associations. The ‘known binary’ associations constituted a smaller percentage of total associations identified in each cohort as we moved from L0 to L3 cohorts, consistent with our expectations that the case-prediction signals may diminish as a function of relaxing the MILTON probability inclusion threshold (Figure 5A, Supplementary Fig. 5C).

**Figure 5.**
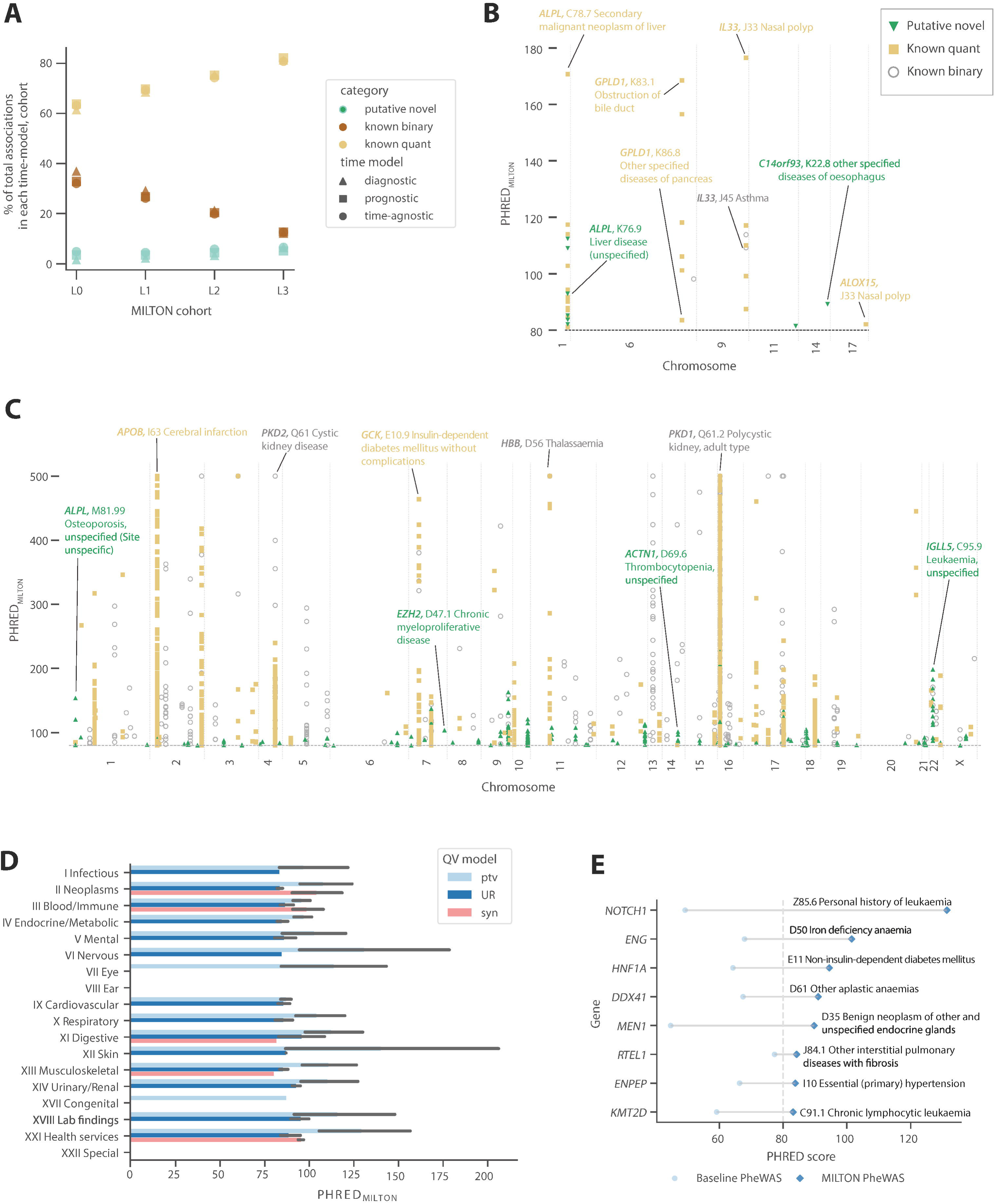
**A** Distribution of known binary (p_MILTON_<1×10^−8^ + p_Baseline_< 1×10^−8^), known quant (p_MILTON_<1×10^−8^ + p_Baseline_> 1×10^−8^+ p_quant PheWAS for FIS>1.5_<1×10^−8^) and putative novel (p_MILTON_<1×10^−8^ + p_Baseline_> 1×10^−8^ + p_quant PheWAS for FIS>1.5_>1×10^−8^) associations across multiple MILTON augmented cohorts and time-models. **B-C** Manhattan plots showing the distribution of gene-ICD10 associations with OR<1 (**B**) and OR>1 (**C**) across different chromosome positions. **D** Distribution of PHRED scores from putative novel (p<10^−8^) gene-disease associations by chapter and across 3 out of the 11 QV models employed by MILTON. “ptv”: protein truncating variant, “UR”: ultra-rare variant, “syn”: synonymous variant. More details about these models can be found in Wang et al. 2021^2^. Error-bars denote 95% confidence intervals and end of the bars represent mean PHRED scores per chapter. **E** Examples of known gene-disease associations from literature that reached genome-wide significance via MILTON.

**Table 2:**
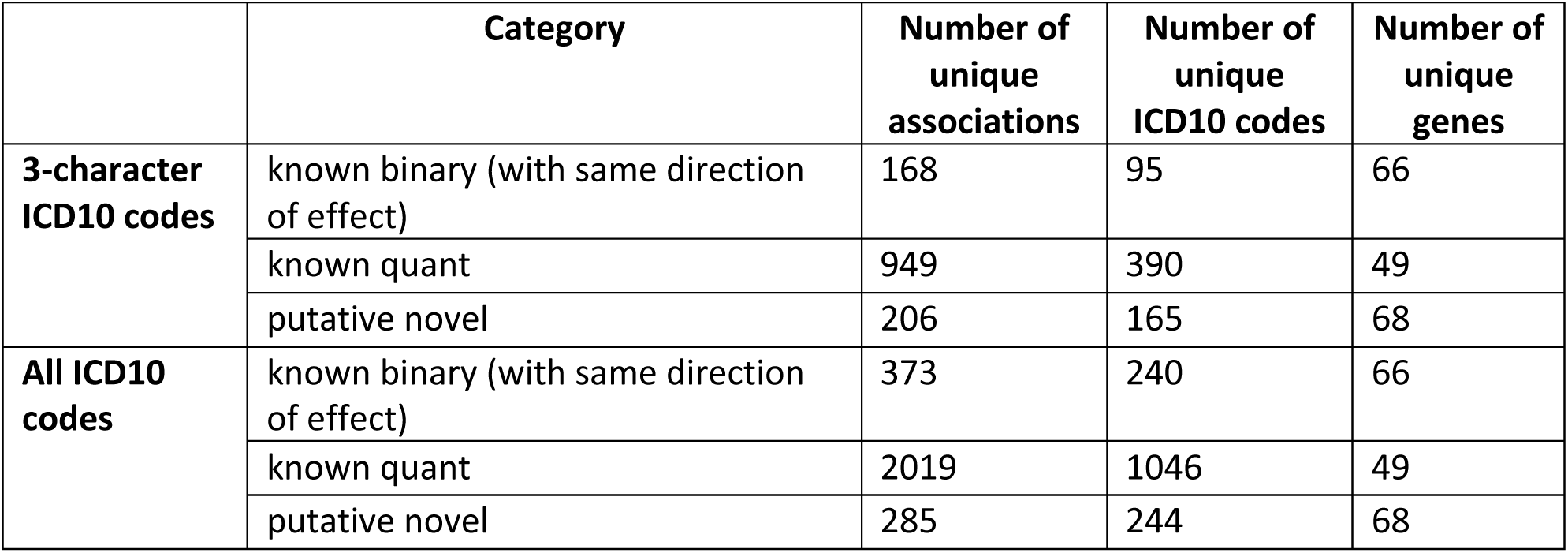
Number of unique gene-ICD10 associations (p<1×10^−8^). The gene-ICD10 association with lowest p-value across all 10 non-synonymous QV models, 3 time-models and 4 augmented cohorts was reported.

We then investigated the effect of highly predictive biomarker feature(s) per ICD10-code on remaining 1,155 gene-ICD10 associations, to distinguish any putative novel signals that may be reflecting more the correlation with a biomarker than specifically disease contribution by comparing to genes achieving significance with that biomarker in the baseline quantitative PheWAS. These features could dominate the prediction of new cases in a way that genes associated with these quantitative traits become associated with the overall ICD10 in PheWAS. To identify these associations, we first shortlisted feature(s) per ICD10-code that had FIS greater than 1.5 standard deviations away from the mean FIS per ICD10-code (see Methods and Materials; Supplementary Fig. 5D). We, therefore, labelled all genes associated with a given ICD10-code that were also significantly associated with its highly predictive feature(s) in quantitative PheWAS results from Wang et al. 2021^2^ (p_quant PheWAS for FIS>1.5_<1×10^−8^) as “known quant”. These accounted for 949 significant gene-disease associations (p_MILTON_<1×10^−8^ & p_Baseline_> 1×10^−8^ & p_quant PheWAS for FIS>1.5_<1×10^−8^; Table 2).

Finally, we labelled the remaining 206 gene-disease associations as ‘putative novel’ (p_MILTON_<1×10^−8^ & p_Baseline_> 1×10^−8^ & p_quant PheWAS for FIS>1.5_>1×10^−8^; Table 2). Please note that the 206 putative novel associations refer to 3-character level ICD10 codes, to reduce over-reporting of novel associations across similar diseases. Overall, MILTON reported 285 putative novel hits across all ICD10 codes (Table 2), 58.7% of which also reached nominal significance (p<0.05) in baseline PheWAS. These putative novel associations were not identified when PheWAS analysis was performed on known cases augmented with randomly selected individuals (see Methods and Materials). The distribution of these putative novel associations by chromosome position (Figure 5B, 5C) and ICD10 chapters are shown in Supplementary Fig. 5E. As a positive control observation, Chapters XXI (Health services) and XVIII (Lab findings) were ranked as the top chapters with putative novel associations, as it inherently refers to individuals who have atypical biomarker levels which is the focus of MILTON (Supplementary Fig. 5E). Importantly, we observed novel associations are predominantly predicted for the non-synonymous QV models (Figure 5D), representing 97.89% of all putative novel hits reported by MILTON. Some of the positive controls^23–37^ for which MILTON enhanced PheWAS results are shown in Figure 5E. For example, the association of protein truncating variants (ptv) in regulator of telomere elongation helicase 1 (*RTEL1*) gene with other interstitial pulmonary diseases with fibrosis (J84.1)^34,35^ achieved p-value of 1.85 × 10^−8^ in baseline cohort but a p-value of 3.75×10^−9^ in MILTON L0 cohort under time-agnostic model. Compared to baseline, the number of cases with ptv QVs increased from 14 to 22 in the L0 cohort (the increase in statistical power is a function of a larger case sample since we also observe the change in case carrier frequency decreases from 0.55% to 0.37%) while number of controls with ptv QVs decreased from 214 to 236 (change in control carrier frequency had a more modest reduction due to larger sampling and went from 0.0734% to 0.0733%). Similarly, the association of protein truncating variants (ptv) in multiple endocrine neoplasia type 1 (*MEN1*) gene with D35 benign neoplasm of other and unspecified endocrine glands^30,38^ had a p-value of 3.3×10^−5^ in baseline cohort but a p-value of 1.06×10^−9^ in L2 cohort under prognostic time-model. The total number of D35 cases used for rare-variant collapsing analysis increased from 1,722 to 8,294 in L2 cohort with an increase in cases with ptv in *MEN1* from 3 (0.17% carrier frequency) to 7 (0.08% carrier frequency) and decrease in controls with ptv in *MEN1* from 10 (0.003%) to 4 (0.001%).

### Genes identified via rare-variant collapsing analyses on MILTON augmented cohorts also rank highly in Mantis-ML 2.0 predictions

Mantis-ML^39^ considers orthogonal, publicly available gene-centric resources and knowledge graphs to assign ranks for each gene in the human exome across thousands of phenotypes. Stepwise-hypergeometric tests were performed comparing Mantis-ML 2.0^40^ and associations identified by MILTON (Figure 6A, see Methods and Materials). These tests aim not only to quantify the overlap between the two methods, which can be quantified by a simple Fisher’s exact test, but are rather suited to include the ranking of the results in the assessment of the overlap enrichment. We performed these tests for fourteen of these phenotypes in Mantis-ML 2.0 where a mapping between human phenotype ontology^41^ (HPO) and ICD10 was manually identified (Supplementary Table 5).

**Figure 6.**
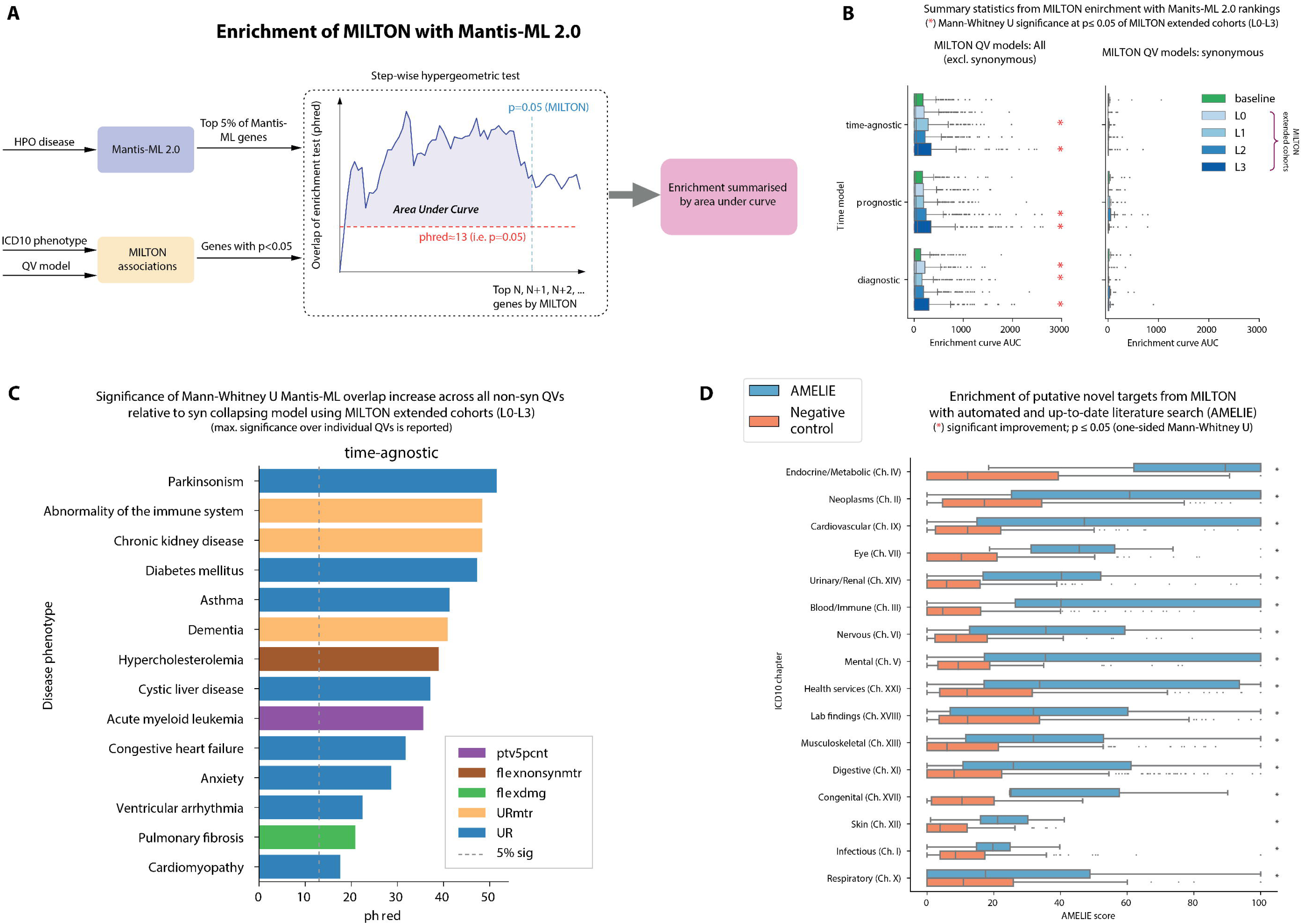
Validation of PheWAS results on MILTON augmented cohorts using two orthogonal methods: Mantis-ML 2.0 and AMELIE. **A** Flowchart explaining the step-wise hypergeometric tests performed to test enrichment (see Methods and Materials). **B** Box plots comparing the enrichment curve AUC between MILTON augmented cohorts and baseline cohorts across all three time-models and non-synonymous QV models on the left panel vs synonymous QV model on the right panel. **C** Bar plot showing the QV model per disease area that achieved the highest phred score (−10log_10_(p-value)) over synonymous QV model. **D** Breakdown of AMELIE aggregated scores by ICD10 chapter (sorted by chapter median) for putative novel targets per three-character ICD10 code. Negative control were generated through 10 samplings of random gene sets, equal in size to the respective MILTON gene sets. In **B** and **D**, each box-plot shows median as centre line, 25^th^ percentile as lower box limit, 75^th^ percentile as upper box-limit, whiskers extend to 25^th^ percentile – 1.5 * interquartile range at the bottom and 75^th^ percentile + 1.5*interquartile range at the top, points denote outliers.

Interestingly, genes identified in L3 cohorts for non-synonymous QV models (p<0.05) were significantly enriched in top-ranking genes from Mantis-ML 2.0 compared to baseline cohort across all three time-models (Figure 6B; comparison per phenotype is shown in Supplementary Fig. 6A). L3 is the largest of all augmented cohorts and contributed the highest number of putative novel associations across all four cohorts (Figure 5A and Supplementary Fig. 5C).

Furthermore, all phenotypes were enriched in top-ranking genes under ultra-rare damaging QV model (UR; UKB minor allele frequency ≤0.005%^2^) in MILTON augmented cohorts compared to synonymous QV model, followed by UR variants according to missense tolerance ratio (URmtr^2^) model and protein truncating variant (ptv^2^) model (Supplementary Fig. 6B). These validations pinpoint MILTON’s power to highlight putative novel signals also supported by comprehensive biological evidence in literature and dozens of public data resources.

### Putative novel targets have higher enrichment with up-to-date literature compared to random genes

As an additional assessment, we performed an automated literature search using AMELIE^42,43^ (version 3.1.0), integrating nightly updates from the entire PubMED corpus, to estimate gene rankings for disease causality among a candidate gene pool. As part of the evaluation process, AMELIE requires diseases to be organised within the Human Phenotype Ontology (HPO). As such, it is necessary to exploit natural language processing (NLP) techniques to fully automate the procedure across a diverse range of disease-gene associations. More specifically, we found the 5 most semantically similar HPO diseases out of a pool of 17,451 phenotypes to each ICD10 code (at the three character level) and queried AMELIE for disease-gene associations for those diseases. We calculated semantic similarity through taking the Euclidean distance between the average BioWordVec^44^ embeddings per disease term in the ICD10 and HPO disease terms.

The results of AMELIE for a given HPO-gene pair is a score (out of 100) representing the gene ranking. This is returned for up to 5 PubMed articles per disease-gene pair. We therefore aggregated (taking the mean) over these 5 articles to provide a single summary statistic of the rank of a disease-gene pair – taking into account that there are 5 HPO diseases per ICD10 code (found using NLP). We observed that for a given ICD10 chapter, putative novel targets (Supplementary Data 4) had significantly higher AMELIE score (p<0.05) than randomly selected targets of same length (Figure 6D). The top 10 phenotypes per chapter by AMELIE enrichment scores are shown in Supplementary Fig. 7.

## Discussion

We have presented here a machine-learning framework called “MILTON” for empowering case-control studies. MILTON achieved median AUC > 0.7 across 953 ICD10 codes and can also successfully identify individuals who subsequently received a diagnosis. MILTON also identified a collection of predictive biomarkers per ICD10-code within a 10-year window of sample collection and pushed 1,155 gene-disease associations beyond genome-wide significance threshold (p<1×10^−8^) in binary PheWAS. By using quantitative PheWAS results from Wang et al. 2021^2^, we were able to further disentangle 949 gene-disease associations possibly dominated by the most predictive feature learnt by MILTON per disease. Moreover, our nominally significant (p<0.05) and putative novel associations (p<1×10^−8^) were preferentially enriched among Mantis-ML^39,40^ and AMELIE^42,43^ predicted gene-disease associations, as two independent sources of additional evidence.

In some occasions, applying MILTON reduced the power for signal discovery by diluting gene-disease signals from the baseline analysis that was naïve to MILTON. However, overall, 78.5% of the known signals from baseline PheWAS either got an improved signal or retained their genome-wide significance with same direction of effect. An additional 206 were novel gene-disease signals that were not discovered by either the baseline case-control or quantitative trait PHEWAS. Thus, MILTON more often leads to improved signal detection and discovery yield than diminished signals, providing strong confidence that this approach is truly augmenting the traditional (baseline) association tests relying solely on positively labelled case definitions in large biobanks. Future implementation of MILTON can include flexible time-lag, time-model selection for each ICD10-code as some phenotypes can take less than 10 years to manifest themselves.

MILTON provides a way to enhance rare-variant collapsing analysis while bridging the gap between clinical biomarkers and diseases. The MILTON-inferred biomarker sets may provide insights into defining minimal sets of biomarkers, irrespective of diagnosis, to be collected as part of future Biobanks, and make them available to registered researchers.

## Methods and materials

### Defining cases

All the subjects diagnosed with a given ICD10-code across 4 different UKB fields were considered as cases. These UKB fields are 41270 Diagnosis – ICD10; 40001 Underlying (primary) cause of death: ICD10; 40002 Contributory (secondary) causes of death: ICD10; and 40006 Type of cancer: ICD10. Out of 13,942 ICD10 codes, 12081 codes already had entries for greater than 50% of cases in the field id 41270 (Supplementary Fig. 8A). When analysing a parent node such as N18, patients diagnosed with any of its sub-nodes: N18.0, N18.1, N18.2, N18.3, N18.4, N18.5, N18.8, N18.9 were included as well. If after taking a union, there were at least 100 known cases for each ICD10-code, that ICD10-code was processed further. The following additional filtering steps were applied to these cases when applicable.

### Filtering based on lag between biomarker sample collection and diagnosis

To assess the impact of time lag between biomarker sample collection and diagnosis dates, cases were filtered based on given (time-model, time-lag) pairs:

1. (Time-agnostic, no lag): include all cases irrespective of any time difference between biomarker sample collection and diagnosis.
2. (Prognostic, ‘t’ years lag): only include those cases who received diagnosis between 0 years and ‘t’ years (inclusive) after biomarker sample collection.
3. (Diagnostic, ‘t’ years lag): only include those cases who received diagnosis between 0 years and ‘t’ years (inclusive) before biomarker sample collection.

Therefore, if time-model=prognostic and time-lag=5 years, then MILTON pipeline will be trained on 85% of all known cases who were diagnosed ≤ 5 years of biomarker collection. In case of multiple lag values per subject, the shortest time-lag was used for analysis. This will happen when analysing a parent node such as ‘N18’ and the patient has already been diagnosed with its sub-nodes such as N18.1 and N18.3 at two different time-points. As the diagnosis information can be retrieved from any of the 4 UKB fields (41270, 40001, 40002 and 40006), the corresponding date field was used to retrieve time for diagnosis: date of first in-patient diagnosis (41280), date of death (40000) and date of cancer diagnosis (40005).

Please note that the time-lag was calculated as difference between biomarker collection date (or biomarker sample collection date in case biomarker was measured from blood or urine samples) and diagnosis date. This difference was converted to days and divided by 365 to obtain time-lag in years.

We performed a sensitivity analysis for the effect of the sample collection and diagnosis time-lag on 400 randomly selected ICD10 codes. Within each time-model, no significant difference in performance was observed across time-lags (two-sided Mann-Whitney U test: p>0.05, Supplementary Fig. 1A). Furthermore, diagnostic models had consistently higher performance than prognostic models across all time-lags tested (two-sided Mann-Whitney U test: p<0.05, Supplementary Fig. 1A). This difference remained consistent across all ICD10-chapters (Supplementary Fig. 1B) and increased with increasing time-lag (Supplementary Fig. 1C). However, for time-lags >= 5 years, diagnostic models had significantly lower cases for training than prognostic models (two-sided Mann-Whitney U test: p≤1×10^−3^, Supplementary Fig. 1D). Additionally, the number of cases available for training increased with increasing time-lag (Supplementary Fig. 1D). This is also evident from the lag distribution across all UKB individuals irrespective of specific diagnosis (Figure 2A). A longer time-lag resulted in more ICD10 codes satisfying the minimum 100-cases criterion for training MILTON models (see Methods and Materials). Therefore, we selected 10 years as the optimal time-lag since: a) median performance of prognostic models started dropping after the 10 years’ time-lag, b) it had comparable performance with other time-lags for diagnostic models, and c) the number of cases attained with a 10 years’ time-lag were significantly higher compared to the 1 or 5 years’ time-lags (Supplementary Fig. 1D).

### Filtering based on sex-specificity of an ICD10-code

Some diseases might affect the opposite sex in rare occasions or not at all given anatomical differences between males and females. To have sufficiently clean data for training a machine learning model, cases from the opposite sex were filtered out if an ICD10-code is deemed to dominantly affect one sex. To find a suitable threshold for filtering, 117 male-specific and 606 female-specific ICD10 codes were shortlisted by keyword search. For male-specific diseases, keywords: “male”, “prostate”, “ testi” and “patern” keywords were used. For female-specific diseases, keywords: “female”, “breast”, “ovar”, “uter”, “pregnan” and ‘’matern” were used. Only those ICD10 codes with at least 100 cases in total were considered for this analysis resulting in 31 male-specific and 201 female-specific ICD10 codes (Supplementary Table 6). As shown in Supplementary Fig. 8B, 231 out of 232 diseases had proportion of dominant sex above 0.9 with highest number of females = 12 for N40 Hyperplasia of prostate (#males=26,033) and highest number of males = 128 for C50.9 Breast, unspecified (#females=15,311). The ICD10-code N62: Hypertrophy of breast (#males=317, #females=864) did not satisfy the ±0.9 threshold and includes a condition called Gynaecomastia that causes abnormal enlargement of breasts in males. Therefore, N62 was not considered to be a female-specific phenotype in our analysis.

### Defining controls

All the remaining UKB subjects who were not diagnosed with any of the ICD10 codes in the entire Chapter to which the current ICD10-code belongs to were considered as ‘potential controls’. To maintain similar case to control ratio across different ICD10 codes, the number of controls was restricted to be four times the number of cases. These ‘controls’ were randomly selected from the list of ‘potential controls’ and following filtering was further applied when applicable.

### Filtering to match baseline characteristics of cases

According to the UKB field ID 31: Sex, there are 54.40% females (n=273,326) and 45.60% males (n=229,086) across the entire UKB cohorts. This imbalance was significant (p-value<1×10^−3^) across multiple age-groups (Supplementary Fig. 8C) as well as multiple ICD10 codes with varying case numbers (Supplementary Fig. 8D). In order to maintain similar distribution of age (UKB field 21003) and sex (UKB field 31) across cases and controls, age was divided into 5 bins. Across these 5 bins, the distribution of females (and males) in controls was matched with the distribution of females (and males) in cases. For example, if 10 females and 5 males were diagnosed with a given ICD10-code and their distribution across 5 hypothetical age bins is given in Supplementary Table 7, then the controls were sampled to match the case distribution as given in Supplementary Table 7. This criterion meant that sometimes cases were dropped to get the matching distribution between cases and controls.

### Features

For each subject, quantitative traits (p=67) collected from blood assays (p=50), urine assays (p=4), physical measures (p=10) along with covariates fasting time, sex and age were used as features for training the machine learning models (Supplementary Table 4).

If a feature was collected during multiple assessment visits (instances 0-3 in UKB), only the first non-null values for each trait were used. The correlation between these features is shown in Supplementary Fig. 9. LDL direct was highly correlated with apolipoprotein B (Pearson’s correlation coefficient = 0.96) and cholesterol (Pearson’s correlation coefficient = 0.95). Similarly, HDL cholesterol was highly correlated with apolipoprotein A (Pearson’s correlation coefficient = 0.92) but negatively correlated with triglycerides (Pearson’s correlation coefficient = −0.44) and waist circumference (Pearson’s correlation coefficient = −0.48).

Despite taking first non-null value across multiple instances, features rheumatoid factor and oestradiol had more than 80% values missing (Supplementary Fig. 10). It has been recommended by UKB to treat these missing values as “naturally low” instead of missing^52^. Further, we observed that the amount of missingness correlated among features obtained from the same assay category, suggesting that these subjects didn’t have the corresponding tests taken (Supplementary Fig. 11). It is unclear why microalbumin in urine had 68.84% missing values while other features obtained from urine assays only had around 3.50% missing values. Its missingness pattern was also similar to that of oestradiol and rheumatoid factor.

Also, blood sample collection dates and urine sample collection dates were recorded in UKB fields 21842 and 21851, respectively. The date for recording physical measures, fasting time, sex and age were assumed to be the date when a participant visited assessment centre (UKB field 53). As the difference between these difference dates is 0 for most of the subject IDs (Supplementary Table 8), blood sample collection dates were used for all features when calculating time-lag between sample collection date and diagnosis date (see Methods and Materials section “Filtering based on lag between biomarker sample collection and diagnosis”).

### Feature pre-processing

The following pre-processing steps were applied on the training data and then on the testing data:

1. Features with 100% missing values were removed;
2. Missing values in each feature (except rheumatoid factor, oestradiol and testosterone) were imputed with median value of that feature (Supplementary Table 4). As it is recommended to treat missing values in rheumatoid factor and oestradiol as “naturally low” instead of missing^52^:

a. Rheumatoid factor was imputed with 0 IU/ml (https://www.ouh.nhs.uk/immunology/diagnostic-tests/tests-catalogue/rheumatoid-factor.aspx) and
b. oestradiol was imputed with values 36.71pmol/L for males and 110.13 pmol/L for females (https://www.southtees.nhs.uk/services/pathology/tests/oestradiol/).
c. Also, testosterone feature was imputed with respective median values for males and females.
3. Categorical features were converted to binary;
4. All features were standardized to have zero mean and unit variance.

### Data preparation

For classification model development, cases were assigned label ‘1’ and controls were assigned label ‘0’. This matched case/control data for each ICD10 code was further randomly divided into training and testing, where 85% of the data was used for training a classification model while remaining 15% of the data was used for validating the performance of the trained model. Cases to controls ratio was preserved between training and testing data.

### Model training

The training of eXtreme Gradient Boosting (XGBoost) classifier was divided into two steps: hyperparameter tuning and model prediction.

#### Hyperparameter tuning

The number of XGBoost trees (‘n_estimators’) were tuned for each ICD10-code. For this, equal number of age-gender matched controls as cases were shortlisted for each ICD10-code. The XGBoost classifier was then fit on the entire data using each of the n_estimators values {5, 15, 50, 100, 200} and its performance was recorded in terms of area under the receiver operating characteristic curve (AUROC). The n_estimators value giving the highest AUROC for each ICD10-code was then used for further analysis.

#### Model prediction

Once the optimal number of n_estimators was determined for each ICD10-code, the next step was to predict the probability of each UKB subject for being a case given their biomarker profile. A predicted probability value closer to 1 denotes higher chances of a subject being a case while a value closer to 0 denotes higher chances of being a control. To obtain these predicted probabilities, steps outlined in the paragraph below were repeated 10 times and average prediction probability across these 10 iterations was calculated for each subject in the entire UKB.

In each iteration, all the cases (*n*) diagnosed with that ICD10-code were taken (see Materials and Methods section “Defining cases”) and controls (4 x *n*) were randomly sampled (see Materials and Methods section “Defining controls”). Five-fold cross validation (CV) was performed where in each fold, four-fifths of the data (cases = *n*\*4/5, controls = *n*\*16/5) was used for training a ‘Balanced Ensemble Classifier’ and remaining one-fifth (cases = *n*\*1/5, controls = *n*\*4/5) was used for testing model performance. A Balanced Ensemble Classifier trains a XGBoost classifier on equal number of cases (*n*\*4/5) and controls (*n*\*4/5) by random subsampling from *n*\*16/5 controls without replacement and repeats the training process 4 times to include all controls. The average performance from these 4 Balanced Ensemble Classifier repeats is reported as the performance of each CV fold and the average performance across all 5 folds is reported as the performance of each overall iteration. An XGBoost classifier is then fitted on the entire training dataset (cases=*n*, controls=4*n*) to generate prediction probabilities for all UKB subjects.

### MILTON case cohort generation

For each ICD10-code, 4 novel case ratio (NCR)-based cohorts, namely ‘L0’, ‘L1’, ‘L2’ and ‘L3’ were generated. Here, NCR is defined as 1 + [(number of new cases)/(number of known cases)]. The distribution of sex for each new cohort was matched with that of known cases.

To generate NCR-based MILTON cohorts with known and new cases for each ICD10-code, all known cases were assigned an average prediction probability of 1 and any not-known subject with probability less than 0.7 was filtered out. All the remaining not-known subjects were ranked in the decreasing order of their average prediction probability, i.e. a subject with highest prediction probability will have rank 1, second highest prediction probability will have rank 2 and so on. These ranks were then divided by the number of known cases to obtain a (NCR – 1) score per subject. Therefore, if the total number of known cases is 10, then adding all not-known subjects with rank less than or equal to 3 will give an NCR of 1+(3/10) = 1.3. The range of NCR was clipped between 1 (no new case) and 10 (number of new cases = 9 x number of known cases) and it was divided into four quantiles: NCR_q1_, NCR_q2_, NCR_q3_, NCR_q4_. All not-known cases with rank less than equal to (NCR_q1_ − 1)*(number of known cases) was added to ‘L0’ cohort, all not-known cases with rank less than equal to (NCR_q2_ − 1)*(number of known cases) was added to ‘L1’ cohort and so on for ‘L2’ and ‘L3’ cohorts. All known cases were then added to each of these NCR-based case cohorts.

### PheWAS collapsing analysis on case cohorts

The PheWAS collapsing analysis was performed on each (ICD10 code, case cohort, qualifying variant (QV) model) combination using whole exome sequencing data from 405,703 UKB participants of European ancestry^2^. To do this, all the non-cases according to each (ICD10-code, case cohort) pair were taken as controls and matched with sex distribution of cases. A 2 x 2 contingency table was generated between cases and controls with or without qualifying variants (QVs) in each gene^2^. Fisher’s exact test with alternative “greater” was performed with the null hypothesis that there is no difference in the number of QVs in each gene between cases and controls^2^. All genes with a p-value less than 1×10^−8^ were considered to be significantly associated with the given ICD10 code for a given QV model and case-control cohort.

Clonal haematopoiesis (CH) is the presence of clonal somatic mutations in the blood of individuals without a haematologic malignancy^53^, and these somatic variants can be detected with germline variant callers^54^. To avoid conflating somatic with germline variants, and age confounded disease associations, we removed from further analysis fifteen genes we previously identified in the UK Biobank as carrying clonal somatic mutations^48^ (*ASXL1, BRAF, DNMT3A, GNB1, IDH2, JAK2, KRAS, MPL, NRAS, PPM1D, PRPF8, SF3B1, SRSF2, TET2, TP53*). A few other problematic genes as listed in Wang et al. 2021^2^ have also been removed from further investigation.

### Feature importance score calculation

During model training, XGBoost classifier was enabled to return the relative biomarker (feature) importance scores (FIS) for each feature. FIS was calculated for each ‘n_estimator’ in XGBoost using Gini purity index that quantifies performance improvement when a given feature is used for making decision split. This process was repeated for all ‘n_estimators’ and average FIS scores were reported for each trained XGBoost model. Because we used a BalancedEnsembleClassifier for training 4 different XGBoost models on 4 different subsamples of controls (see Methods and Materials section “Model prediction”), average of these 4 average relative FIS scores were reported for each ICD10-code for each feature.

### Finding optimal feature importance score cut-off

For each time-model, non-zero feature importance scores (FIS) per ICD10-code were first log-transformed and then standardised to have zero mean and unit standard deviation. Top FIS per ICD10-code per time-model was extracted and compared with the median AUC of ICD10-code. It was observed that FIS of top features highly correlated with AUC performance of ICD10 codes (Pearson’s correlation coefficient > 0.80; Extended Fig. 4B). This suggested that high performing models with AUC > 0.80 identified dominating features and assigned high feature importance scores to them (>4 standard deviations away from the mean FIS). A threshold of 4 was therefore chosen for further analysis.

### Comparison with Mantis-ML 2.0

Stepwise-hypergeometric tests proceed through identifying a ranked list of genes (ordered from most to least associated according to Mantis-ML 2.0^39,40^ and a set of genes ‘known’ to be associated. The latter may be a list of tentatively associated genes, provided e.g. by a PheWAS with a relaxed p-value threshold. The test then iteratively goes through the ranked list marking the N, N+1, N+2 etc genes as associated in the ranked list and quantifies the overlap between this and the latter (the set of known associated genes) using Fisher’s exact test. In this way we may observe whether there is substantial overlap between the two without specifying a threshold for the ranked list. The output of this exercise is a sequence of p-values that can be converted to PHRED scores using −10log10(p). Finally, to provide a simple summary statistic for the test we may take the area under the curve (AUC), where the curve is defined as max(−10log10(0.05), PHRED) – i.e. the area exceeding 0.05 significance. Requiring that the curves exceed 0.05 significance allows a certain degree of noise to be filtered out. In this setting, genes from MILTON with a weak significance were used and the top 5% of Mantis-ML 2.0 associated genes were used as the ranked list.

Please note that Mantis-ML 2.0 uses human phenotype ontology (HPO), and MILTON uses ICD10 codes and manual HPO to ICD10 mapping was performed (Supplementary Table 5).

### Comorbidity enrichment analysis

The enrichment analysis performs a Fisher’s exact test for all distinct ICD10 diagnoses within the selected cohort of UK Biobank subjects to find diseases which have significantly higher incidence in the cohort as compared with the general population (the rest of UK Biobank subjects). ICD10 codes are scored according to the results of their Fisher’s tests to highlight the most relevant ones. Fisher’s exact test produces by default two numbers: p-value and odds ratio. The former is a measure of how significant the disease enrichment in the selected cohort is, while the latter corresponds to the effect size, i.e., how much more likely the subjects from the selected cohort are to be diagnosed with the disease. The score then is taken to be the log of odds ratio, capped at a high value, and normalized to 100 to facilitate visualization of results.

## Data availability

All the biomarker information, diagnosis information along with relevant dates and whole exome sequencing data can be obtained from the UKB (http://www.ukbiobank.ac.uk/register-apply/). The list of 67 quantitative traits along with their UKB field ids is given in Supplementary Table 4 and can be found on the UKB showcase portal (https://biobank.ndph.ox.ac.uk/showcase/search.cgi). Data for this study were obtained under Resource Application Number 26041.

All results produced in this study will be made available on the MILTON web-portal.

## Code availability

MILTON accesses UKB individual level data, therefore, the developed software packages will be made available through the UKB portal.

## Author contributions

Conception and design of the study: D.V and S.P.. Developed the method and generated phenome-wide results: M.K., M.G., and D.V.. Contributed to methods and analytical strategies: M.G., M.K., D.M. L.M., A.O’N., Q.W., A.H., R.D., S.P., and D.V. Developed the web portal: D.M., M.K., and D.V.. Performed validation analyses: L.M., M.G., and M.K.. Contributed to exploration and interpretation of results: M.G., M.K., L.M., J.M., A.H., R.D., S.P., and D.V. Wrote the paper: M.G., L.M., J.M., A.H., R.D., S.P., and D.V. All authors provided input and revisions for the final paper.

## Acknowledgements

We thank the UK Biobank team and participants for making this valuable resource possible. We also thank the AstraZeneca Centre for Genomics Research Analytics and Informatics teams, and Margarete Fabre for their feedback.

