## Supplementary Figures for "Applying Machine Learning on UK Biobank biomarker data empowers case-control discovery yield"

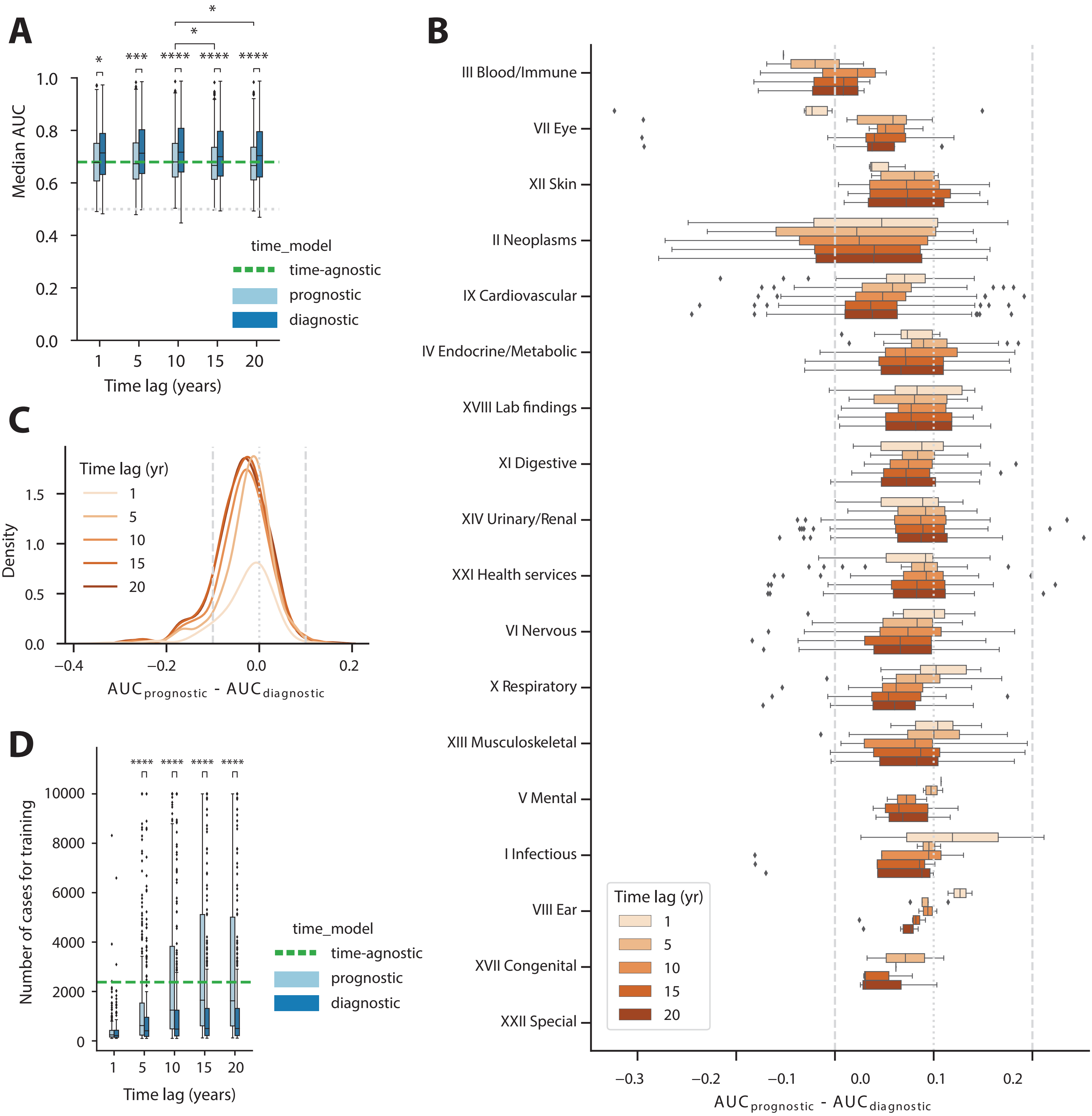


**Supplementary Fig. 1 A** Boxplots comparing the overall performance of MILTON-models across 400 random ICD10 codes when trained on cases satisfying the given time-lag and time-model. Gray, dashed line represent an AUC of 0.5, equivalent to random guesses. Number codes with results for (diagnostic, prognostic) time-models, respectively: 1 year (157, 192); 5 years (323, 371); 10 years (368, 371); 15 years (400, 400); 20 years (400, 399). Time-agnostic model had results for 400 codes. **B** Boxplots comparing the performance of MILTON-models across each ICD10-chapter when trained on cases satisfying the given time-lag and time-model. **C** Density plot showing the distribution of difference between prognostic and diagnostic models across ICD10 codes. **D** Boxplots comparing the number of cases satisfying the given time-lag available for training. Mann-Whitney U test, two-sided p-values are shown in panels A and D. *: 1.00e-02 < p ≤ 5.00e-02; **: 1.00e-03 < p ≤ 1.00e-02; ***: 1.00e-04 < p ≤ 1.00e-03; ****: p ≤ 1.00e-04. In panels A, B and D, the box-plot shows median as centre line, 25$^{th}$ percentile as lower box limit, 75$^{th}$ percentile as upper box-limit, whiskers extend to 25$^{th}$ percentile – 1.5 * interquartile range at the bottom and 75$^{th}$ percentile + 1.5*interquartile range at the top, points denote outliers.

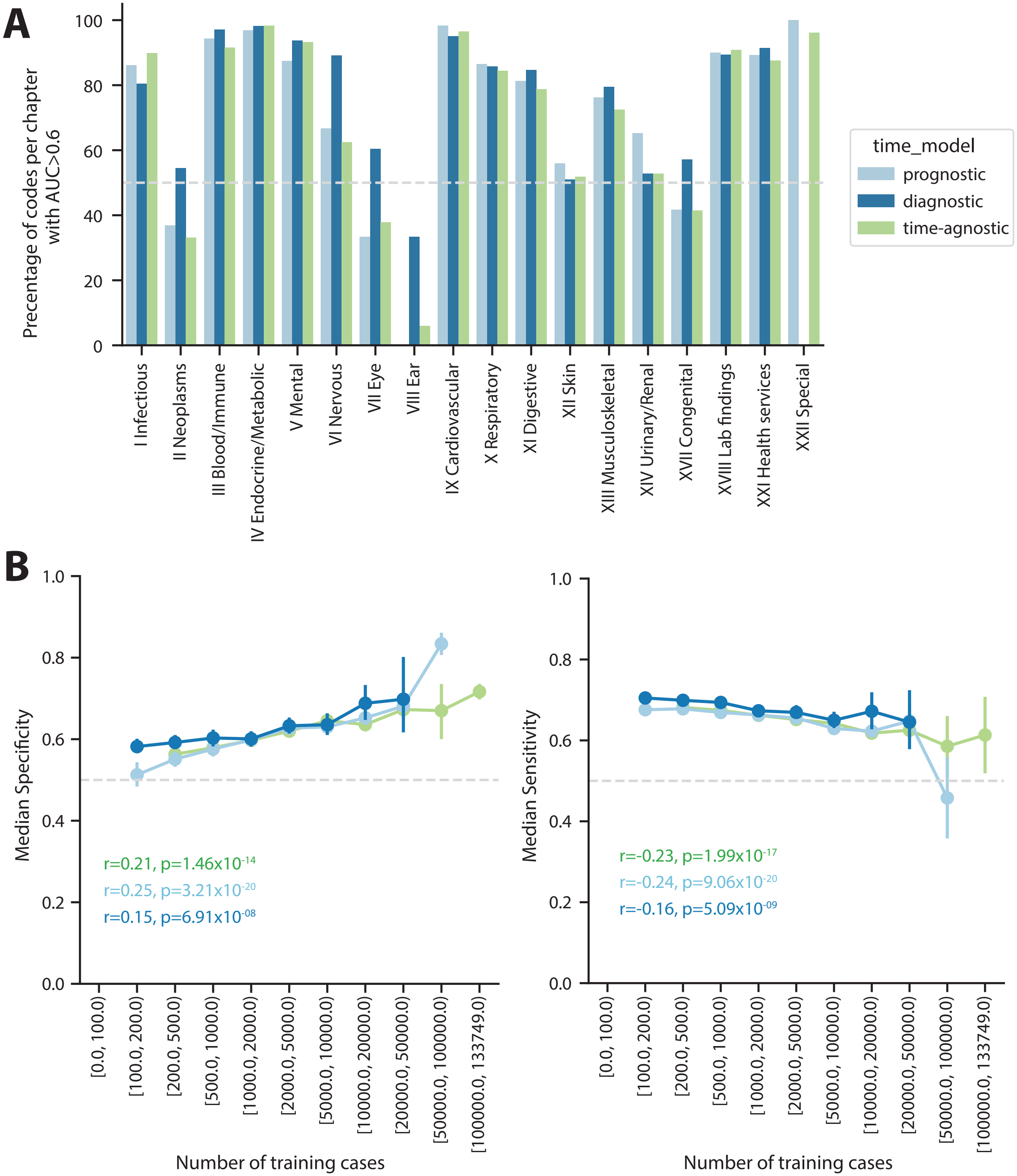


**Supplementary Fig. 2 A** Bar plots comparing the overall performance of MILTON-models across ICD10 chapters. Gray dashed line represents that 50% of ICD10 codes per chapter had AUC > 0.6. **B** Point-plots showing the variation of sensitivity and specificity with increasing number of cases available for training in each ICD10-code. Error-bar represents 95% confidence interval. Pearson correlation coefficients (r) and p-values (p) for each time-model are indicated.

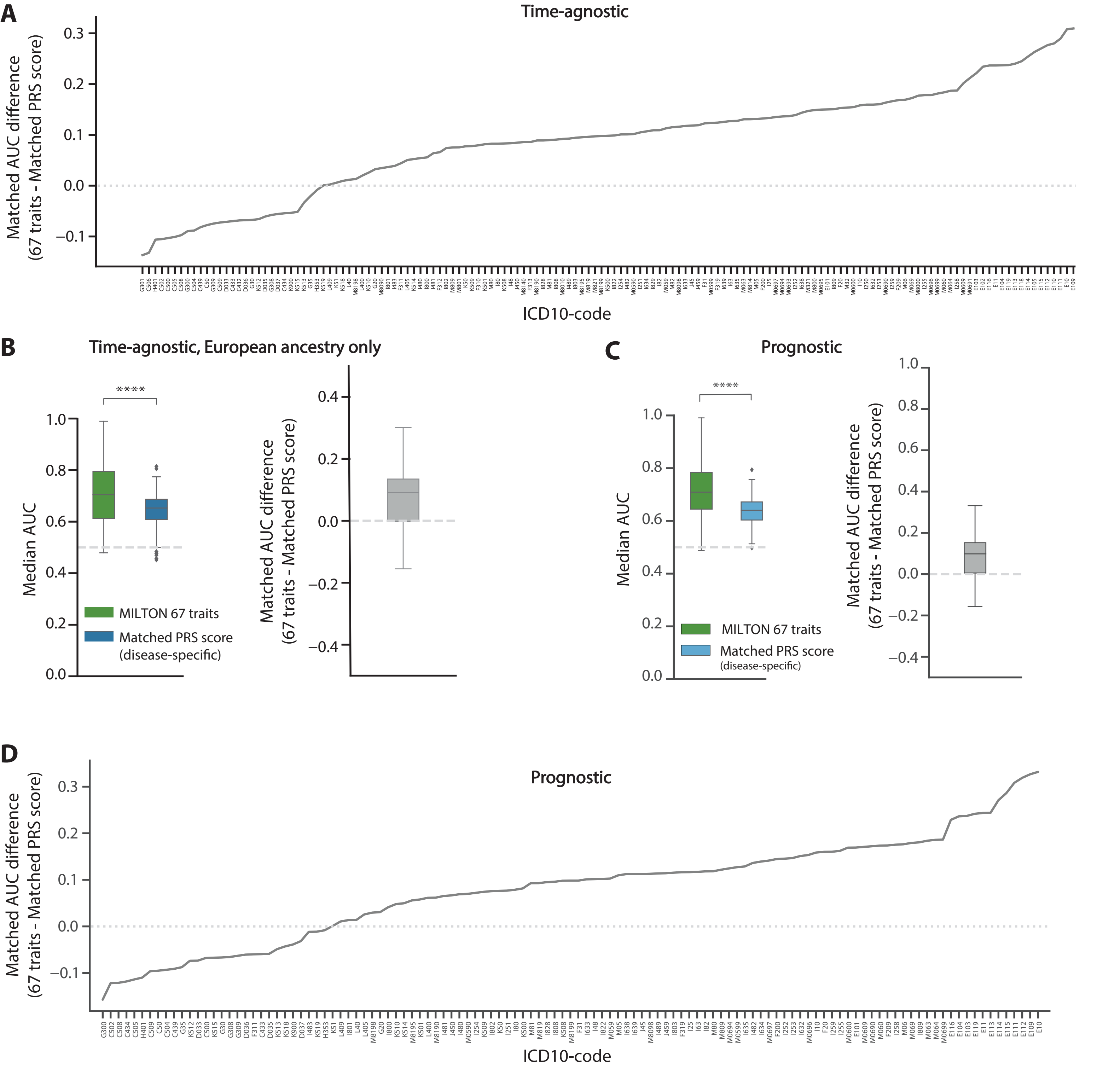


**Supplementary Fig. 3 A** Line-plots comparing the median AUC (y-axis) of MILTON time-agnostic models trained 67 traits with disease-specific PRS scores for each disease (x-axis). **B** Boxplots comparing the performance of MILTON time-agnostic models trained on individuals from European ancestry only using either 67 traits or disease-specific PRS scores across 152 ICD10-codes. **C** Boxplots comparing the performance of MILTON prognostic models trained on 67 traits with disease-specific PRS scores. **** Mann-Whitney U test, two-sided p-values < $10^{-4}$. Right panel: x-axis represents Median $AUC_{67\ traits}$ – Median $AUC_{Disease\text{-}specific\ PRS}$. In panels B and C, each box-plot shows median as centre line, 25$^{th}$ percentile as lower box limit, 75$^{th}$ percentile as upper box-limit, whiskers extend to 25$^{th}$ percentile – 1.5 * interquartile range at the bottom and 75$^{th}$ percentile + 1.5*interquartile range at the top, points denote outliers. **D** Line-plots comparing the median AUC (y-axis) of MILTON prognostic models trained 67 traits with disease-specific PRS scores for each disease (x-axis).

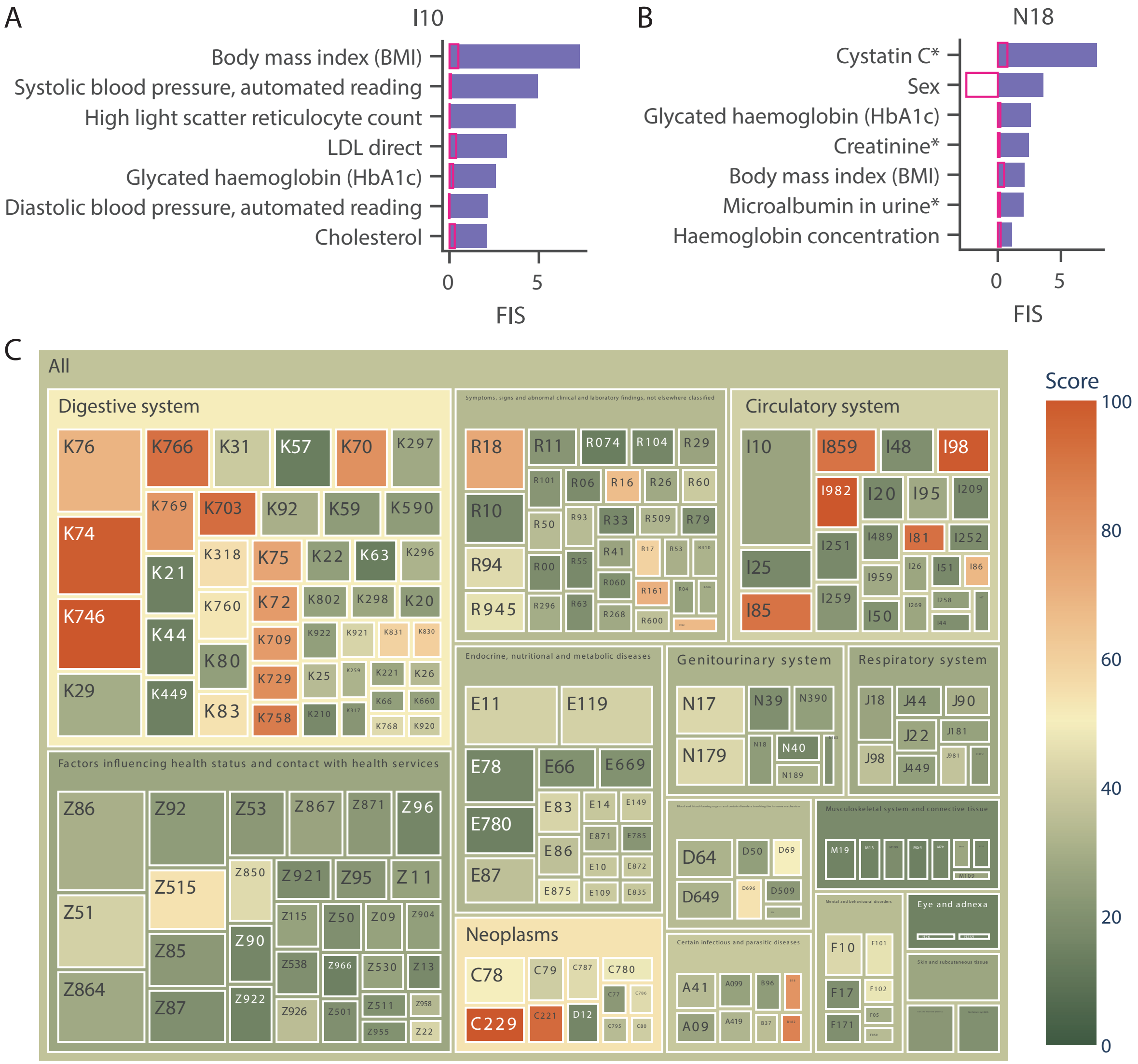


**Supplementary Fig. 4 A-B** Bar plots showing top seven features by standardized feature importance scores (FIS) for each ICD10-code for time-agnostic model. Median FIS for each feature and time-agnostic model across all ICD10 codes is shown in pink bar-plots. I10: Essential (primary) hypertension; J33: Nasal polyp. *: represents biomarkers mapped to the given therapy area by clinical experts[14]. **C** Treemap showing comorbidities in 425 patients diagnosed with C22.0 in UKB. The ICD10 codes with higher log odds ratio (score) compared to other ICD10 codes within each chapter are shown in dark red color.

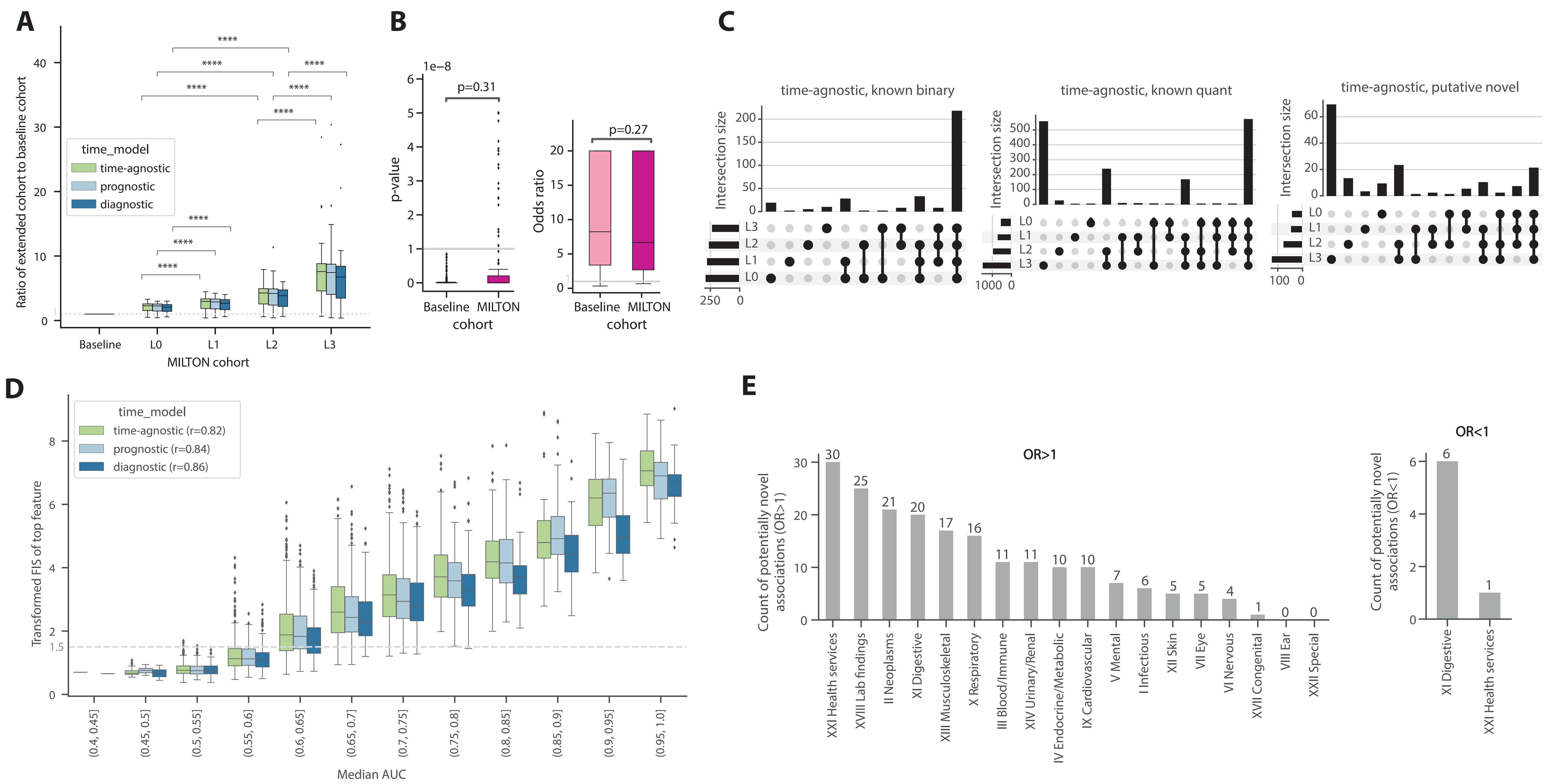


**Supplementary Fig. 5 A** Box plots comparing the cohort ratio [(number of cases in baseline cohort + number of newly predicted cases)/number of cases in baseline cohort] across each MILTON cohort. Baseline cohort has a cohort ratio of 1. ****: Mann-Whitney U test two-sided $p < 10^{-4}$ are shown. **B** Box plot comparing the p-values (left) and odds ratio (right) of PheWAS performed on baseline cohorts used for training MILTON models with MILTON extended cohorts. Mann-Whitney U test two-sided p-value are shown. **C** UpSet plots showing the distribution of known binary, known quant and putative novel gene-ICD10 associations across different MILTON extended cohorts for time-agnostic model. **D** Box plot comparing the standardized FIS of top feature per ICD10-code with the median AUC value of that ICD10-code. Dotted line represents standardized FIS=4. Pearson's correlation coefficient (r) is indicated in brackets in-front of each time-model in the legend. **E** Bar plot showing the number of unique gene-ICD10 associations per chapter. In panels A, B and D, each box-plot shows median as centre line, $25^{th}$ percentile as lower box limit, $75^{th}$ percentile as upper box-limit, whiskers extend to $25^{th}$ percentile – 1.5 * interquartile range at the bottom and $75^{th}$ percentile + 1.5*interquartile range at the top, points denote outliers.

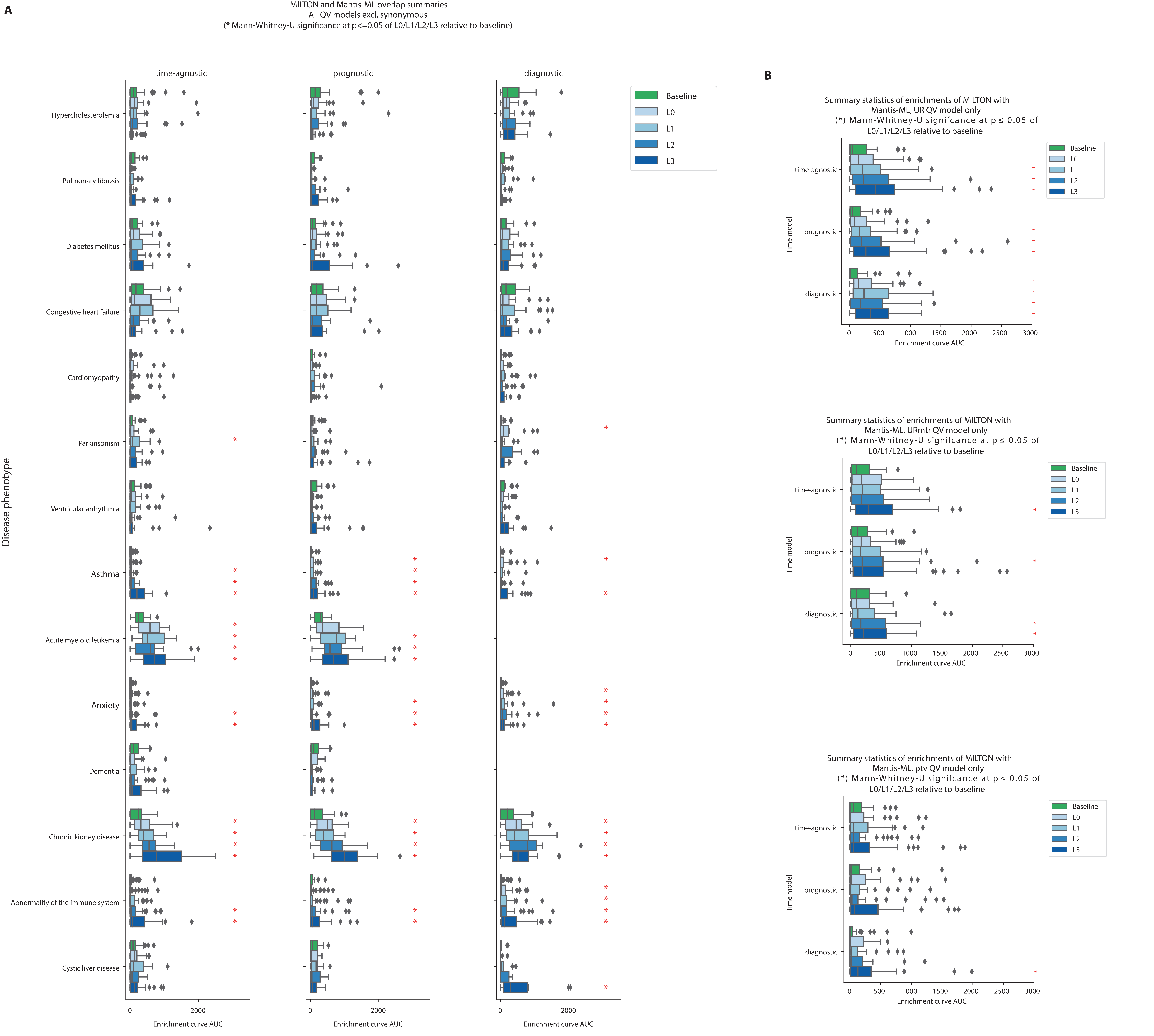


**Supplementary Fig. 6** Enrichment of high-ranking MANTIS-ML genes in MILTON extended cohorts (PheWAS p<0.05) compared to baseline cohorts. **A** Per disease-area and non-synonymous QV models. **B** All disease areas and UR, URmtr and ptv QV models. More details about these QV models can be found in Wang et al. 2021[2]. In panels A and B, each box-plot shows median as centre line, 25[th] percentile as lower box limit, 75[th] percentile as upper box-limit, whiskers extend to 25[th] percentile – 1.5 * interquartile range at the bottom and 75[th] percentile + 1.5*interquartile range at the top, points denote outliers.

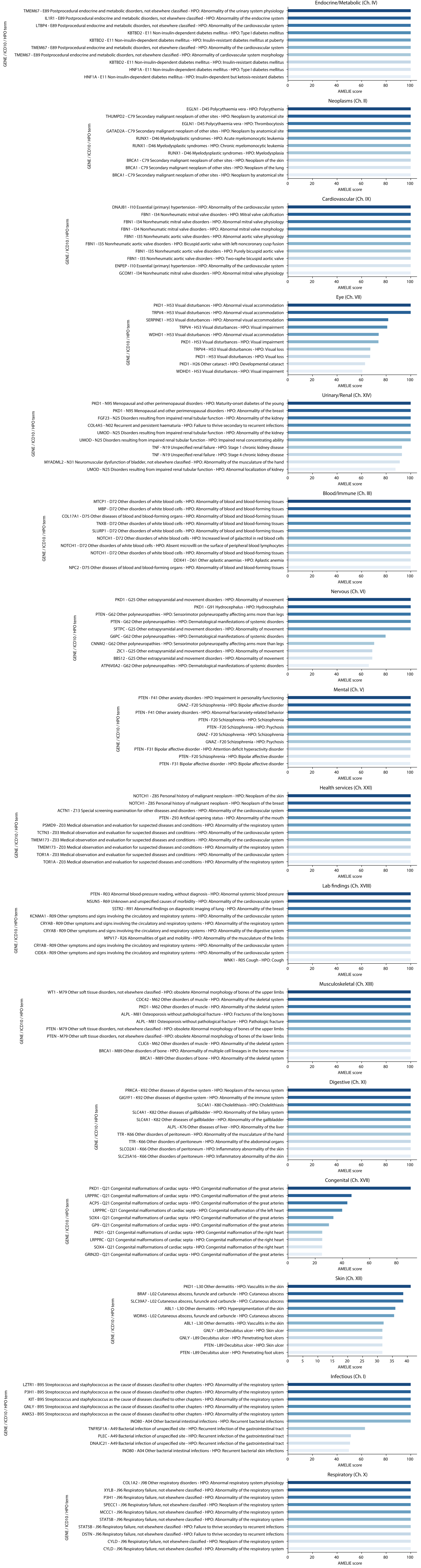


**Supplementary Fig. 7** Bar plots showing top 10 phenotypes by AMELIE enrichment scores (x-axis) for each chapter. For each y-axis label, putative novel gene-ICD10 (at 3-character level) association is shown followed by one of the 5 HPO terms this ICD10 term was mapped to using natural language processing.

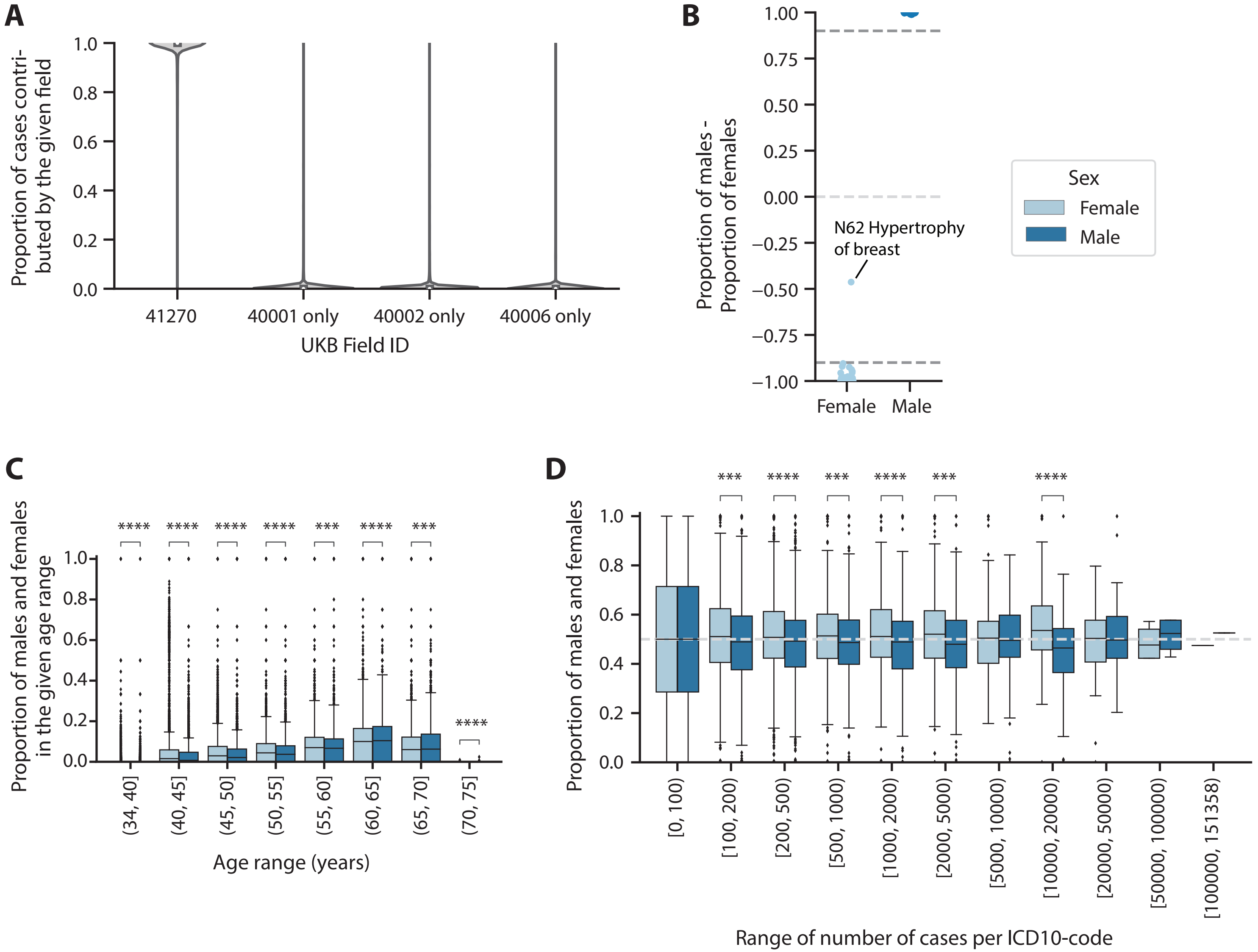


**Supplementary Fig. 8** **A** Violin plot showing the proportion of diagnosed cases contributed by UKB fields 41270: hospital inpatient diagnosis, 40001: underlying (primary) cause of death, 40002: contributory (secondary) causes of death and 40006: Cancer registry. **B** Scatter plot showing the difference in proportion of males and females in sex-specific diseases with value of –1 expected for female-specific diseases and value of 1 expected for male-specific diseases. Dotted lines indicate proportion of ±0.1. **C** Boxplots showing the distribution of males and females in different age-ranges. **D** Boxplots showing the distribution of males and females across ICD10 codes with increasing number of diagnosed cases. Mann-Whitney U test, two-sided p-values are shown in panels C and D. ns: $p > 5.00\text{e-}02$, *: $1.00\text{e-}02 < p \leq 5.00\text{e-}02$; **: $1.00\text{e-}03 < p \leq 1.00\text{e-}02$; ***: $1.00\text{e-}04 < p \leq 1.00\text{e-}03$; ****: $p \leq 1.00\text{e-}04$. In panels C and D, each box-plot shows median as centre line, $25^{th}$ percentile as lower box limit, $75^{th}$ percentile as upper box-limit, whiskers extend to $25^{th}$ percentile – 1.5 * interquartile range at the bottom and $75^{th}$ percentile + 1.5*interquartile range at the top, points denote outliers.

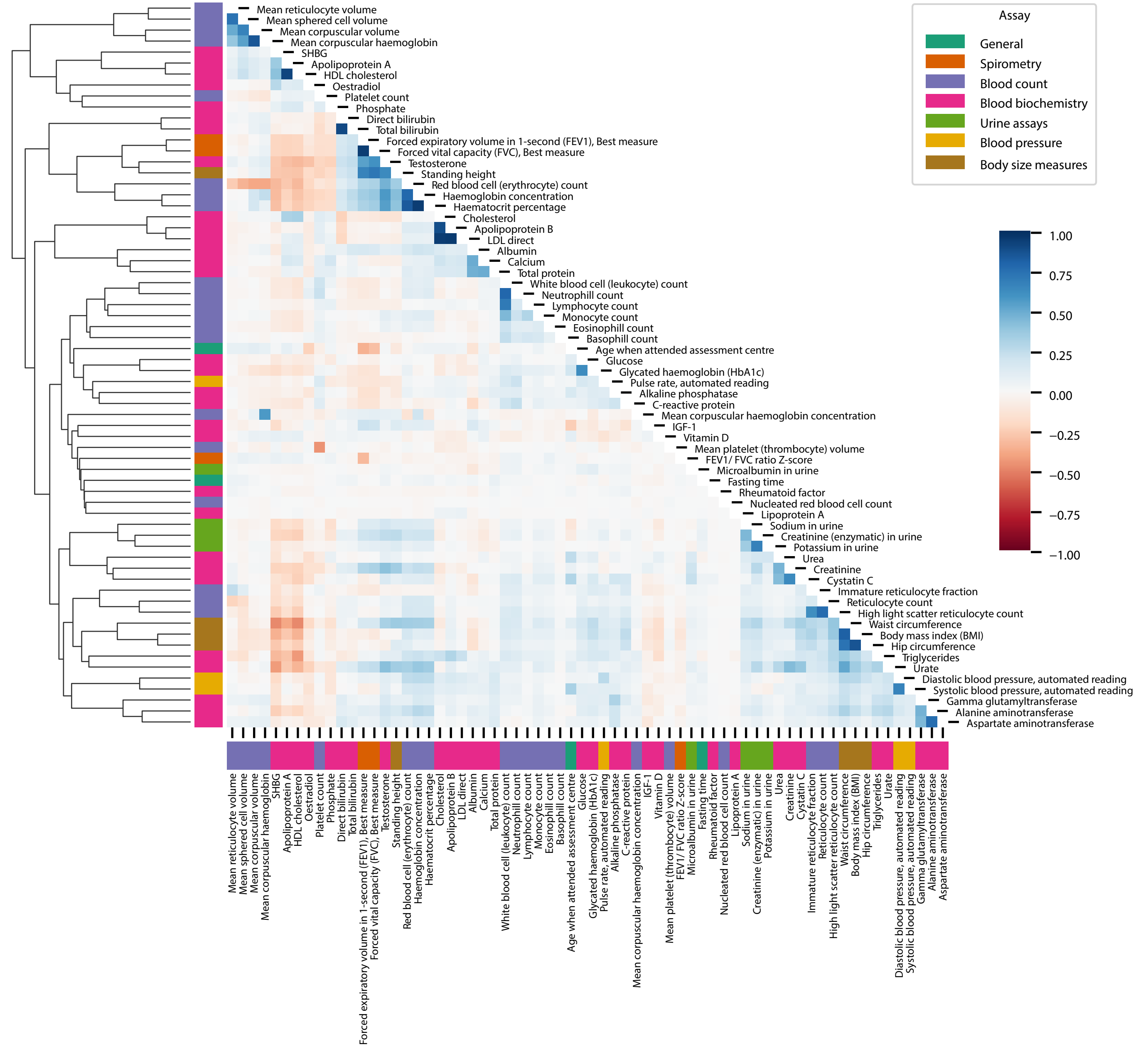


**Supplementary Fig. 9** Heatmap showing Pearson's correlation coefficient between non-missing values in features.

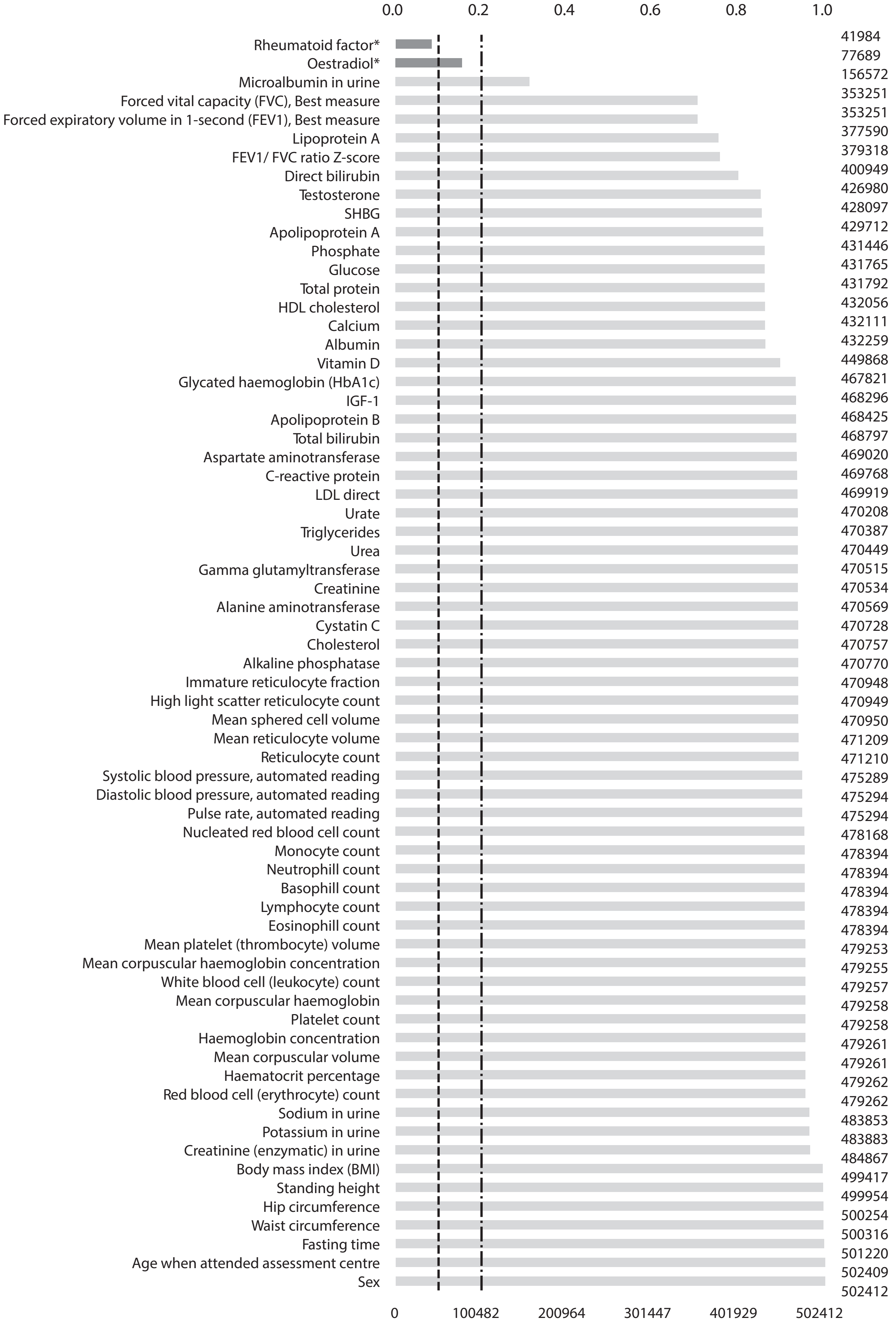


**Supplementary Fig. 10** Bar plot showing the proportion (top) and number of UKB individuals (right) with non-missing values for a given feature (left). * indicates features with greater than 80% missing values.

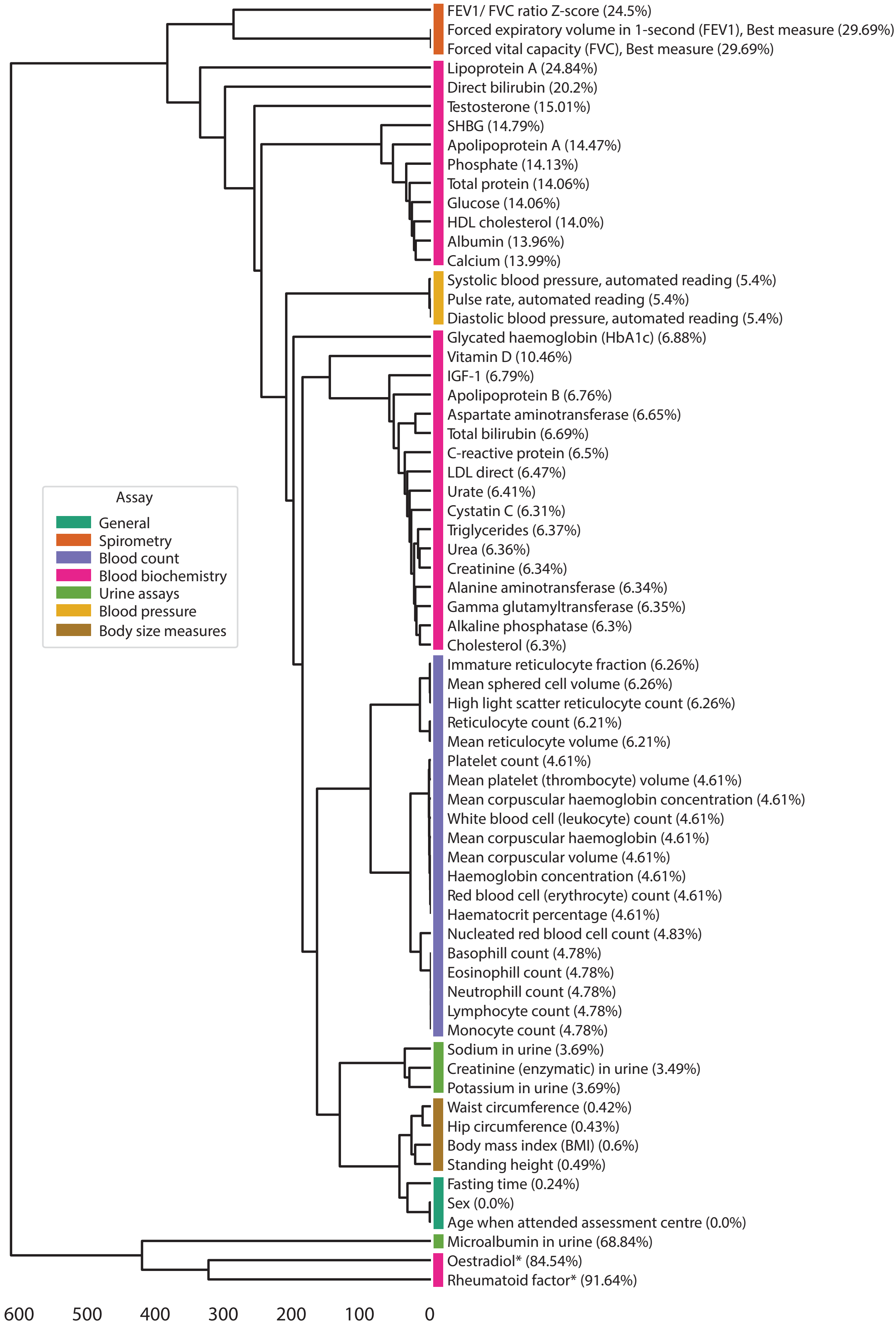


**Supplementary Fig. 11** Dendrogram clustering the features based on their missingness pattern with amount of missingness indicated in brackets. * indicates features with more than 80% missing values.

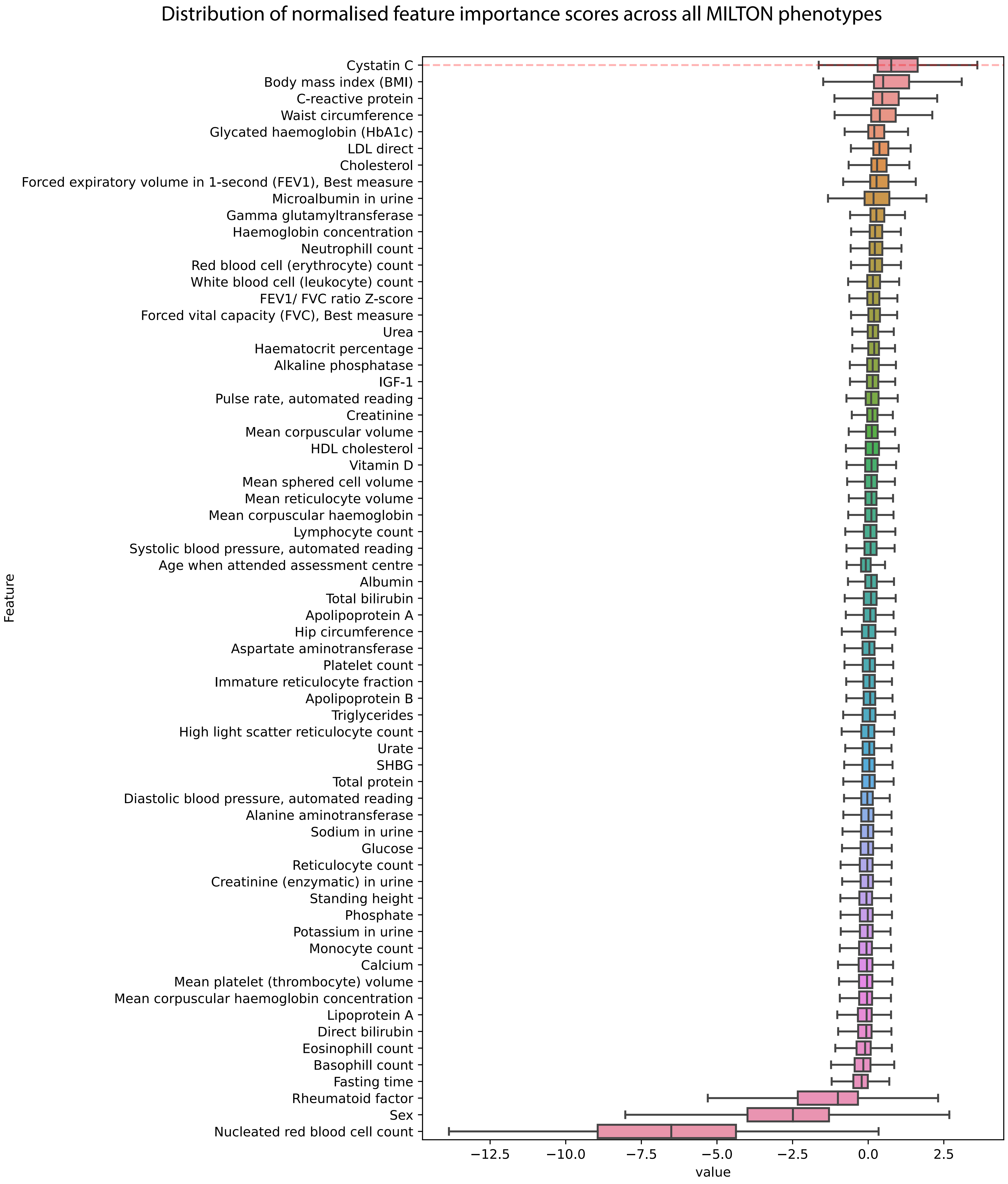

Distribution of normalised feature importance scores across all MILTON phenotypes
Cystatin C
Body mass index (BMI)
C-reactive protein
Waist circumference
Glycated haemoglobin (HbA1c)
LDL direct
Cholesterol
Forced expiratory volume in 1-second (FEV1), Best measure
Microalbumin in urine
Gamma glutamyltransferase
Haemoglobin concentration
Neutrophill count
Red blood cell (erythrocyte) count
White blood cell (leukocyte) count
FEV1/ FVC ratio Z-score
Forced vital capacity (FVC), Best measure
Urea
Haematocrit percentage
Alkaline phosphatase
IGF-1
Pulse rate, automated reading
Creatinine
Mean corpuscular volume
HDL cholesterol
Vitamin D
Mean sphered cell volume
Mean reticulocyte volume
Mean corpuscular haemoglobin
Lymphocyte count
Systolic blood pressure, automated reading
Age when attended assessment centre
Albumin
Total bilirubin
Apolipoprotein A
Hip circumference
Aspartate aminotransferase
Platelet count
Immature reticulocyte fraction
Apolipoprotein B
Triglycerides
High light scatter reticulocyte count
Urate
SHBG
Total protein
Diastolic blood pressure, automated reading
Alanine aminotransferase
Sodium in urine
Glucose
Reticulocyte count
Creatinine (enzymatic) in urine
Standing height
Phosphate
Potassium in urine
Monocyte count
Calcium
Mean platelet (thrombocyte) volume
Mean corpuscular haemoglobin concentration
Lipoprotein A
Direct bilirubin
Eosinophill count
Basophill count
Fasting time
Rheumatoid factor
Sex
Nucleated red blood cell count
Feature
−12.5
−10.0
−7.5
−5.0
−2.5
0.0
2.5
value
